# Multi-ancestry meta-analysis identifies 2 novel loci associated with ischemic stroke and reveals heterogeneity of effects between sexes and ancestries

**DOI:** 10.1101/2022.02.28.22271647

**Authors:** Ida Surakka, Kuan-Han Wu, Whitney Hornsby, Brooke N. Wolford, Fred Shen, Wei Zhou, Jennifer E. Huffman, Anita Pandit, Yao Hu, Ben Brumpton, Anne Heidi Skogholt, Maiken E. Gabrielsen, Robin G. Walters, The TOPMed Stroke Working Group, Million Veteran Program (MVP), Kristian Hveem, Charles Kooperberg, Sebastian Zöllner, Peter W.F. Wilson, Nadia R. Sutton, Mark J. Daly, Benjamin M. Neale, Cristen J. Willer, the Global Biobank Meta-analysis Initiative (GBMI)

## Abstract

Cerebrovascular accident (stroke) is the second leading cause of death and disability worldwide. Stroke prevalence varies by sex and ancestry, which could be due to genetic heterogeneity between subgroups. We performed a genome-wide meta-analysis of 16 biobanks across multiple ancestries to study the genetic contributions underlying ischemic stroke (60,176 cases, 1,310,725 controls) as part of the Global Biobank Meta-analysis Initiative (GBMI). Two novel loci associated ischemic stroke with plausible candidate genes, *FGF5* and *CENPQ/MUT*, were identified after replication in four additional datasets. One locus showed significant ancestry heterogeneity (*PDE3A*) and two loci showed significant sex-heterogeneity (*SH3PXD2A* and *ALDH2*). The *ALDH2* locus had a male-specific association for stroke in GBMI (P-value males = 1.67e-24, P-value females = 0.126). To test whether we would see a difference in the predictive power of sex-specific polygenic risk scores (PRSs), we compared the C-indexes for sex-specific and sex-combined PRSs in HUNT dataset. A sex-combined PRS was more successful at predicting stroke cases than a sex-specific PRS, most likely due to more stable effect estimates from the sex-combined summary-statistics. These approaches can be applied to further unravel the genetic underpinnings of stroke and other complex diseases.

## INTRODUCTION

Cerebrovascular accidents (stroke) are the second leading cause of death and disability worldwide due to brain infarction (ischemic stroke) or intracerebral hemorrhage (Katan and Luft, 2018). The former can be further divided into different subgroups, including cardioembolic, large vessel, and small vessel stroke, and the latter into lobar and non-lobar hemorrhagic stroke. Mapping genetic variants associated with stroke has been more challenging than for other homogenous complex diseases, such as coronary artery disease (Nelson et al., 2017) or type 2 diabetes (Mahajan et al., 2018) given that stroke subgroups have different etiologies (Hankey, 2017) and heritability (Bevan et al., 2012).

Thirty-five loci have been identified using genome-wide association study (GWAS) methods (Malik et al., 2018a; Malik et al., 2018b; Woo et al., 2014) despite the complex phenotypic heterogeneity of stroke. These studies consist of sample sizes up to 900,000 (72,000 cases with all-cause stroke) and show that genetic predisposition varies between subgroups. Most known loci are associated with ischemic stroke, likely due to higher prevalence of that subtype (∼80% of the cases), and thus, more power to detect an association (Donkor, 2018). Additionally, stroke prevalence has been shown to differ between populations of different ancestry and sexes (Guzik and Bushnell, 2017), suggesting possible heterogeneity of environmental and/or genetic factors contributing to the risk for stroke.

The predictive power of polygenic risk scores (PRSs) has been evaluated to preemptively identify stroke risk (Abraham et al., 2019; Marston et al., 2021; Rutten-Jacobs et al., 2018). These scores have shown lower predictive power compared to other cardiovascular disease outcomes (Khera et al., 2018; Mars et al., 2020), likely due to phenotypic heterogeneity and limited sample sizes in the discovery cohorts underlying the PRS calculations. Here, we perform a new GWAS as part of the Global Biobank Meta-analysis Initiative (GBMI) to examine genetic variants and test whether the observed associations and PRS for ischemic stroke show either ancestry-or sex-specific effects.

## RESULTS

### Ischemic stroke locus discovery and ancestry heterogeneity

Following discovery and replication stages, we identified two novel loci associated with ischemic stroke. We initially assessed association summary statistics from 16 biobanks with participants from various ancestries (Zhou et al., 2021) (**Figure 1, Supplementary Table 1)** and performed replication in four additional biobanks (**STAR Methods, Table 1, Figures 2A-B, Supplementary Figures 1A-B**). One variant showed a significant replication P-value (*PRDM8/FGF5* locus, replication P-value < 7.1e-3, Bonferroni correction for 7 tested putative novel variants) and in the other locus (*CENPQ/MUT*), the replication results supported the original discovery results (combining results from discovery excluding BioMe and replication showed more significant association than original discovery alone). Both loci also showed suggestive association in a largely independent meta-analysis of stroke (**Supplementary Figures 2A-B**) with association P-values 4×10^−4^ and 2×10^−4^ for rs12509595 (*PRDM8/FGF5)* and rs2501968 (*CENPQ/MUT)*, respectively (Malik *et al*., 2018a). Replication results for 7 variants tested for replication are in **Supplementary Table 2**.

**Figure 1.**
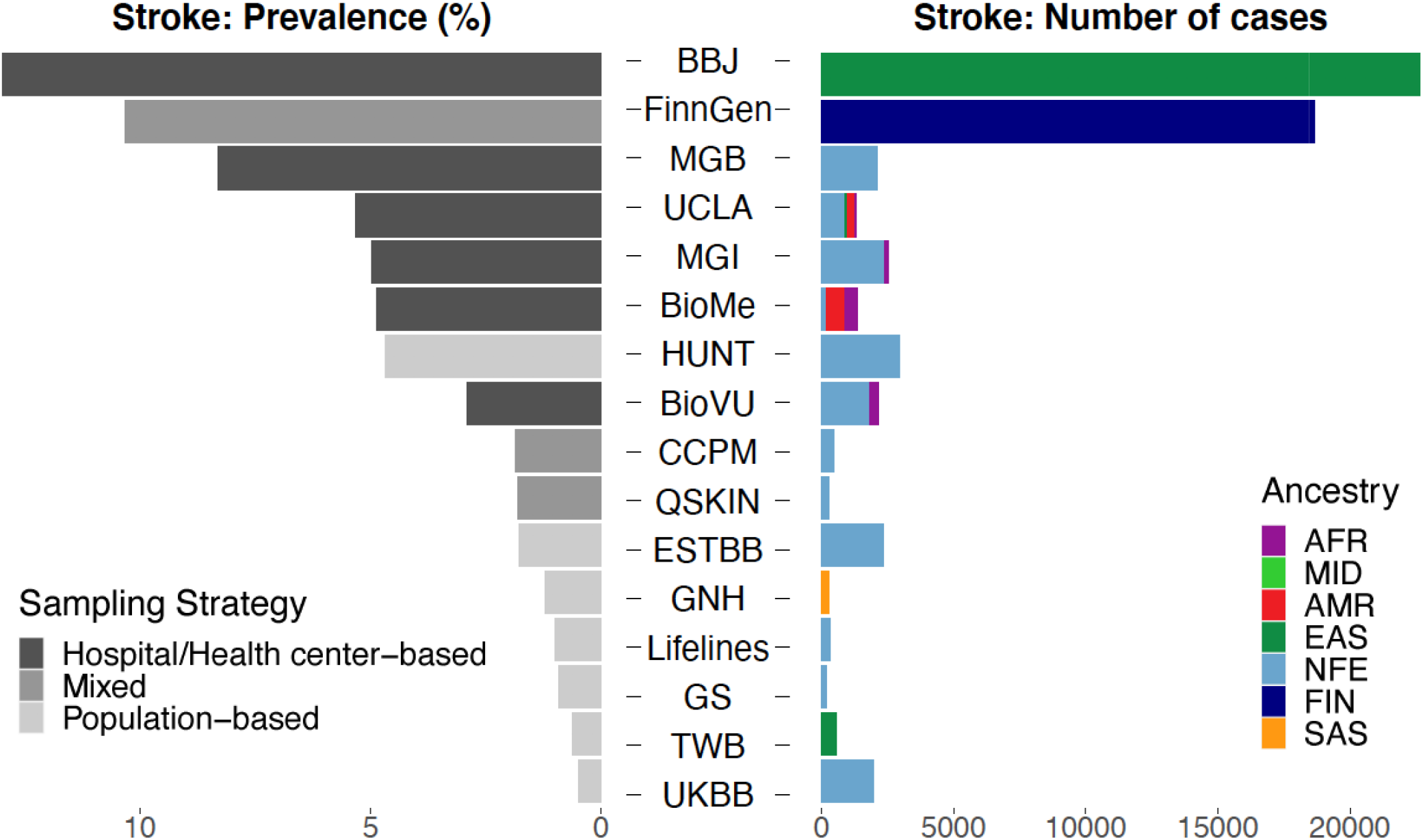
Breakdown of stroke meta-analysis ancestries. This figure presents the prevalence and number of cases by cohort participating in the GBMI stroke meta-analysis. BBJ = Biobank Japan, MGB = Mass General Brigham Biobank, UCLA = UCLA Precision Health Biobank, MGI = Michigan Genomics Initiative, BioMe = Mount Sinai BioMe Biobank, HUNT = Trøndelag Health Study, BioVU = Biorepository at Vanderbilt University, CCPM = Colorado Center for Personalized Medicine, QSKIN = Queensland Skin Study, ESTBB = Estonian Biobank, GNH = East London Genes &Health, GS = Generation Scotland, TWB = Taiwan Biobank, UKBB = UK Biobank, AFR = African ancestry, MID = Middle Eastern ancestry, AMR = Admixed American ancestry, EAS = East Asian ancestry, NFE = non-Finnish European ancestry, FIN = Finnish ancestry, SAS = South Asian ancestry.

**Table 1.**
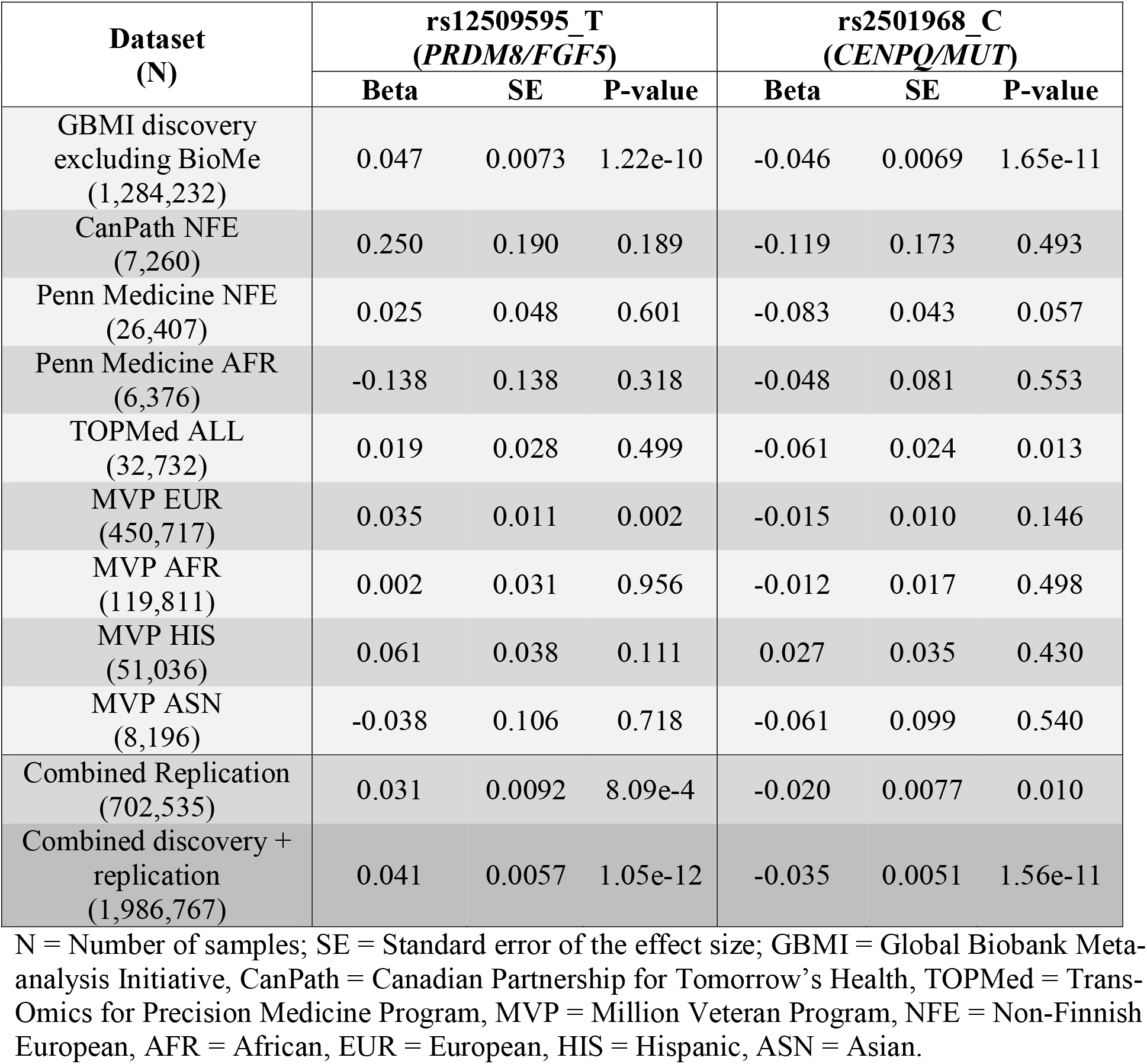
Two newly identified and replicated loci associated with ischemic stroke. For both variants the effect sizes are reported for the minor allele noted also in the legend after the rsID. The discovery results are presented from the leave-BioMe-out summary statistics due to BioME being part of the TOPMed results as well.

**Figure 2.**
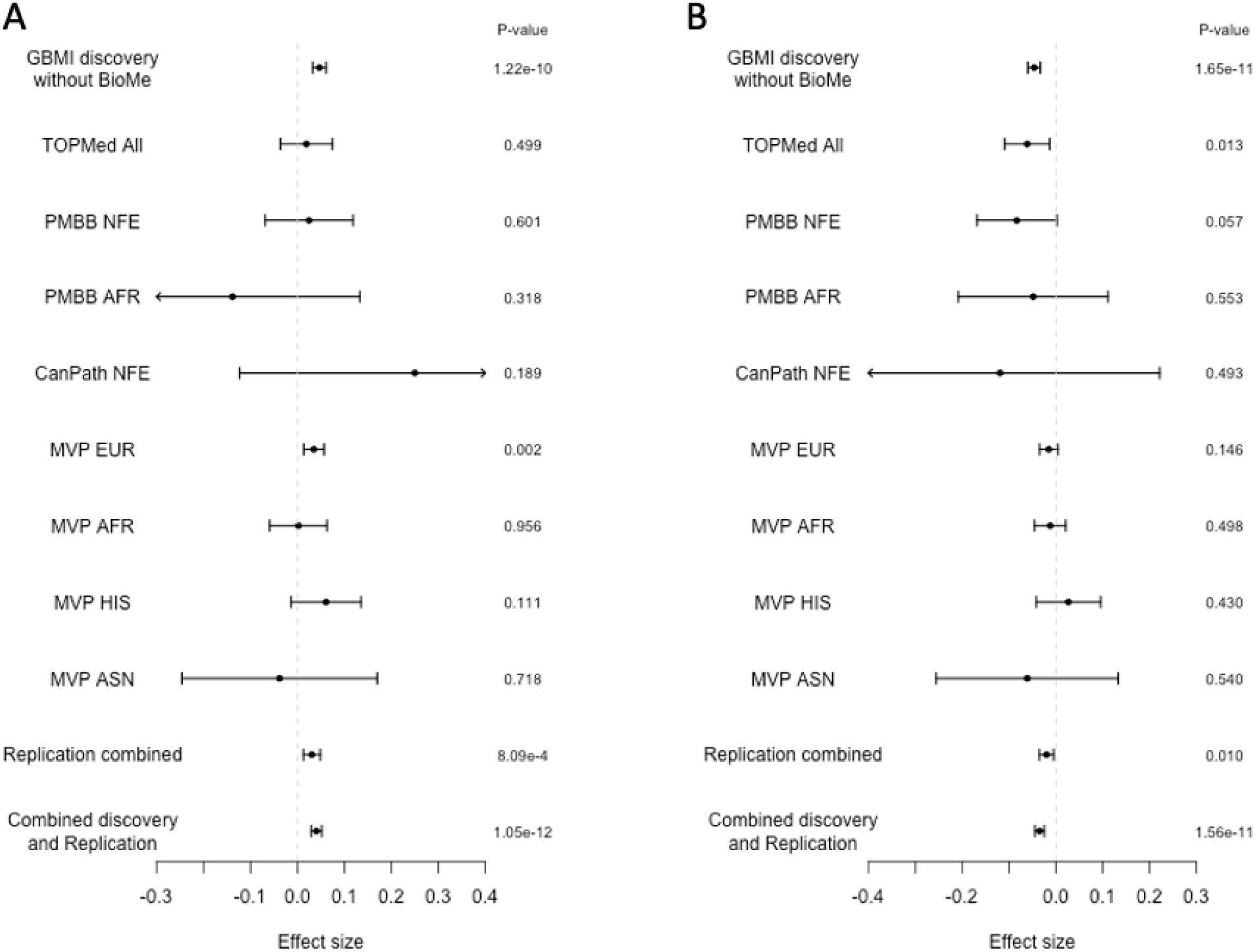
A-B. Discovery and replication results for the two confirmed associations. Presented are the effect sizes together with their 95% confidence intervals. Panel A shows the association results for rs12509595 (*PRDM8/FGF5*) and panel B for rs2501968 (*CENPQ/MUT*). GBMI = Global Biobank Meta-analysis Initiative, BioMe = Mount Sinai BioMe Biobank, TOPMed = Trans-Omics for Precision Medicine Program, PMBB = Penn Medicine Biobank, NFE = Non-Finnish European ancestry, AFR = African ancestry, CanPath = Canadian Partnership for Tomorrow’s Health, MVP = Million Veteran Program, EUR = European ancestry, HIS = Hispanic/Latino ancestry, ASN = Asian ancestry

Of the 2 confirmed novel associations, one index variant was a common missense variant in the gene encoding centromeric protein Q -- *CENPQ* (p.Asp266Gly, rs2501968). However, the index variant was also associated with *CENPQ* gene expression (**Supplementary Table 3A**) in multiple tissues, with the strongest eQTL associations observed in arterial tissues. The variant was also a significant eQTL for the *MUT* gene. Additionally, there were significant splice QTLs for this variant in both CENPQ and MUT genes (**Supplementary Table 3B**). The lead variant for the other confirmed novel association, *PRDM8/FGF5*, was intergenic and showed a significant eQTL for *FGF5* gene expression in the kidney.

Next, the ancestry specific results of the discovered ischemic stroke associations were examined. We observed one locus showing significant ancestry heterogeneity (P-value < 5.5×10^−3^, Bonferroni correction for 9 tests, **Table 2**). The lead variant, rs12811752, lies in the intron of the *PDE3A* gene (a cGMP-inhibited cyclic nucleotide phosphodiesterase) and had consistent effects in the European (non-Finnish European [NFE], and Finns [FIN]), East Asian (EAS), and African (AFR) ancestry populations. However, for the admixed American (AMR) individuals the effect size was approximately 4 times larger. Additionally, for the South Asian (SAS) ancestry individuals the direction of effect was opposite (although non-significant) to that observed in the other ancestry groups. In addition to *PDE3A*, we observed nominally significant ancestry heterogeneity for two loci; *CDKN2B* and *COL4A1*.

**Table 2.** Ancestry specific effect sizes from the GBMI meta-analysis for the significant lead variants. For all variants the effect sizes are reported for the minor allele.

| Variant<br>(Locus) | Effect size<br>(SE) NFE | Effect size<br>(SE) FIN | Effect size<br>(SE) SAS | Effect size<br>(SE) EAS | Effect size<br>(SE) AMR | Effect size<br>(SE) AFR | Cochran's<br>Q-statistic<br>for ancestry | Ancestry<br>heterogeneity<br>P-value |
| --- | --- | --- | --- | --- | --- | --- | --- | --- |
| **Novel variants** |  |  |  |  |  |  |  |  |
| rs12509595<br>(PRDM8/FGF5) | 0.027 (0.014) | 0.037 (0.014) | 0.100 (0.101) | 0.066 (0.011) | 0.142 (0.096) | 0.036 (0.089) | 6.80 | 0.236 |
| rs2501968<br>(CENPQ/MUT) | -0.042 (0.013) | -0.042 (0.013) | -0.043 (0.104) | -0.051 (0.011) | 0.010 (0.064) | -0.082 (0.051) | 1.75 | 0.883 |
| **Previously published variants** |  |  |  |  |  |  |  |  |
| rs1275985<br>(CIB4/KCNK3) | -0.033 (0.013) | -0.032 (0.013) | -0.001 (0.108) | -0.068 (0.013) | -0.055 (0.066) | -0.025 (0.060) | 5.40 | 0.369 |
| rs1333047<br>(CDKN2B-AS1/DMRTA1) | 0.057 (0.013) | 0.091 (0.013) | 0.129 (0.093) | 0.037 (0.011) | 0.111 (0.065) | 0.136 (0.087) | 11.89 | 0.036 |
| rs7091346<br>(SH3PXD2A) | -0.030 (0.013) | NA | -0.130 (0.091) | -0.056 (0.011) | -0.007 (0.064) | -0.134 (0.062) | 5.48 | 0.241 |
| rs12811752<br>(PDE3A) | 0.038 (0.013) | 0.057 (0.013) | -0.065 (0.090) | 0.054 (0.011) | 0.282 (0.041) | 0.040 (0.074) | 34.44 | 1.94e-6 |
| rs671<br>(ALDH2) | -1.040 (3.815) | NA | NA | -0.107 (0.012) | NA | NA | 0.060 | 0.807 |
| rs4773140<br>(COL4A1) | 0.072 (0.016) | 0.007 (0.015) | -0.056 (0.107) | 0.066 (0.012) | -0.078 (0.102) | -0.015 (0.090) | 14.41 | 0.013 |
| rs11880613<br>(DNM2) | -0.054 (0.017) | -0.038 (0.016) | 0.002 (0.152) | -0.058 (0.014) | -0.047 (0.101) | -0.055 (0.134) | 1.065 | 0.957 |

SE = Standard error of the effect size, NFE = non-Finnish European ancestry, FIN = Finnish ancestry, SAS = South Asian ancestry, EAS = East Asian ancestry, AMR = Admixed American ancestry, AFR = African ancestry, NA = Not available

### Sex-specificity of ischemic stroke associations and polygenic risk score

In addition to ancestry heterogeneity, we looked for evidence of sex-specific heterogeneity in the associated loci. Two of the nine associated loci, *ALDH2* and *SH3PXD2A*, showed significant sex-heterogeneity (P-value < 5.56×10^−3^, Bonferroni correction for 9 tests, **Supplementary Table 4, Supplementary Figures 3A-B**). The two significant sex-heterogeneity loci have been associated with stroke in previous GWAS (Malik *et al*., 2018a). The sex-heterogenic effects on ischemic stroke have not been previously reported for *SH3PXD2A*, but have been indirectly implied for *ALDH2* (Millwood et al., 2019). For both loci, the stroke association was stronger in males than for females (effect sizes for males, -0.160 and -0.062, for females -0.031 and -0.031, for *ALDH2* and *SH3PXD2A*, respectively), and significant only in males (P-values for males 1.67e-24 and 4.84e-8, and for females 0.126 and 0.022, for *ALDH2* and *SH3PXD2A*, respectively). At the *ALDH2* locus, the association was East Asian specific, reflecting the absence or very low frequency of this variant in other ancestries in the GBMI meta-analysis. Furthermore, the allele frequency of the lead variant, rs671, was 25.5% for the EAS and ≤0.05% for other ancestries in the publicly available gnomAD database.

We next tested whether a genome-wide polygenic risk score demonstrated sex-differential effects on stroke risk since sex-heterogeneity for multiple lead-variants within the associated loci were observed. Specifically, we created 3 different PRS: for the joint meta-analysis, and for the male- and female-only meta-analyses. The predictive performance of the scores was tested in the HUNT dataset using Cox Proportional Hazards models. The model performance of the joint PRS was low as evidenced by a small change in the model C-index when adding the PRS into the model (C-index for the reference model with age and sex only = 0.858 95% CI [0.849; 0.866], C-index after adding the PRS = 0.860 [0.852; 0.868], **Supplementary Figure 4**). When using the sex-specific PRS instead of the joint PRS, the performance was slightly attenuated for both males (from 0.850 [0.839; 0.862] to 0.848 [0.836; 0.859]) and females (from 0.868 [0.857; 0.880] to 0.867 [0.856; 0.879]), most likely due to decreased power in the sex-specific meta-analysis compared to the joint meta-analysis. Interestingly, when looking at the predictive performance of age only (the reference model), the C-index for females was notably higher (C-index = 0.867 [0.856; 0.879]) compared to males (C-index = 0.846 [0.834; 0.858]). The incremental increase in C-index when adding the ischemic stroke PRS to the prediction model was higher in males, however, this difference between the C-index changes between males and females was not statistically significant.

## DISCUSSION

A genome wide association analysis identified two novel genetic loci associated with stroke, confirmed through replication in four independent datasets. Moreover, we observed significant ancestry heterogeneity in one locus (*PDE3A*) and significant sex heterogeneity in two loci (*ALDH2* and *SH3PXD2A*).

The only protein-altering lead variant resided in *CENPQ*, which has been previously reported to be associated with blood homocysteine levels, a known risk factor for stroke (Paré et al., 2009). *CENPQ* has also been suggested to regulate *MUT* expression, one of the driver genes in the causal relationship between blood homocysteine levels and small vessel stroke (Larsson et al., 2019). The one novel intergenic association was observed near the *FGF5* gene. This locus has been previously associated with blood pressure traits in Chinese individuals with higher body mass index (Li et al., 2015), and the expression of the *FGF5* gene has been shown to be different in hypertensive patients (Ren et al., 2018). Additionally, recent studies have shown that the FGFs could potentially be used to treat stroke in animal models (Dordoe et al., 2021).

Two of the associated loci, rs671 in *ALDH2* and rs7091346 in *SH3PXD2A*, showed significant sex-heterogeneity. The lead variant rs671 is a well-known polymorphism linked to alcohol consumption and hypertension in East Asian individuals (Millwood *et al*., 2019). The effect of this variant is observed more strongly in men, likely due to cultural and societal differences in patterns of alcohol consumption between sexes. We observed an association with ischemic stroke for this variant only in males, and more specifically, in those with East Asian ancestry.

The ischemic stroke PRS did not significantly predict stroke in the HUNT dataset when added on top of the age and sex information. Additionally, in an accompanying paper by Wang et al., the performance of the GBMI derived PRS was compared to one derived using the MegaStroke summary statistics (Wang et al., 2021). Wang et al. concluded that the previous meta-analysis, with more cases underlying the summary statistics, performed better for African ancestry individuals whereas the GBMI derived PRS was slightly better for European individuals. Previous PRS studies have shown that a stroke PRS could slightly increase the prediction of future stroke cases. In a recent study, a stroke PRS derived from the MegaStroke summary statistics performed better than any of the individual risk factors for stroke (Abraham *et al*., 2019). However, they did not evaluate the performance of their score on top of the age and sex as shown here. In our test dataset, we observed a high C-index when using age information only, especially for females (C-index = 0.867 [0.856; 0.879]).

## Limitations

The high representation of East Asian individuals was a notable strength of this investigation, likely leading to the discovery of East Asian driven sex-specific effects in two previously published stroke loci, *ALDH2* and *SH3PXD2A*. Our study has several limitations despite the large overall size of the meta-analysis. First, the ischemic stroke phenotype was defined in the GBMI meta-analyses using an EHR-derived phenotype definition only. This approach does not allow for dissection of results by sub-phenotypes, which has previously been done in large stroke meta-analysis efforts (Malik *et al*., 2018a). Additionally, the number of participating biobanks and total number of individuals for some of the ancestry groups are low (FIN, one biobank N = 180,062; SAS, one biobank N = 21,940; AMR, two biobanks N = 15,064), resulting in limited power to fully test for ancestry-specific effects.

## Outlook/Conclusion

We present 2 novel loci associated with ischemic stroke and show that some stroke loci show sex- and/or ancestry-specific patterns. These findings emphasize the need for more diverse datasets with large enough sample sizes to further understand the genetic predisposition of stroke in different ancestry groups. Finally, we recommend evaluation of polygenic risk scores in males and females separately in different populations, and ideally with stroke subtypes.

## Supporting information

GBMI Author Banner

GBMI Author Acknowledgements

## Data Availability

All data produced in the present study are available upon reasonable request to the authors

## Acknowledgments

We thank Prof. Robin Walters for the critical review. We would like to express our gratitude to all contributors to GBMI and the biobank participants who provided their data for biomedical research. Particularly, we are grateful to GBMI study cohorts, MGI and BioMe, who assisted with replication and interpretation of our findings. The authors acknowledge the participants, recruitment teams and project managers of the GBMI for providing data aggregation, management, and distribution services in support of the research reported in this publication. We acknowledge BioBank Japan (Yukinori Okada, Koichi Matsua, and Masahiro Kanai), BioMe (Ruth Loos, Judy Cho, Eimear Kenny, Michael Preuss, and Simon Lee), BioVU (Nancy Cox and Jibril Hirbo), Canadian Partnership for Tomorrow (Philip Awadalla and Marie-Julie Fave), China Kadoorie (Robin Walters, Kuang Lin, and Iona Millwood), Colorado Center for Personalized Medicine (Kathleen Barnes, Michelle Daya, and Chris Gignoux), deCODE Genetics (Kári Stefánsson and Unnur Þorsteinsdóttir), East London Genes &Health (David A van Heel, Sarah Finer, and Richard Trembath), Estonian Biobank (Andres Metspalu, Reedik Mägi, Tõnu Esko, and Priit Palta), FinnGen (Aarno Palotie, Mark Daly, Samuli Ripatti, Mitja Kurki, and Juha Karjalainen), Generation Scotland (Caroline Hayward and Riccardo Marioni), HUNT (Kristian Hveem, Cristen Willer, and Sarah Graham, Ben Brumpton, and Brooke Wolford), Lifelines (Serena Sanna and Esteban Lopera), Michigan Genomics Initiative (Sebastian Zoellner, Michael Boehnke, Lars Fritsche, and Anita Pandit), Million Veteran Program (Christopher J. O’Donnell), Netherlands Twin Register (Dl Boomsma, MG Nivard), Partners Biobank (Jordan Smoller and Yen-Chen Feng), QIMR Berghofer (Sarah Medland, Stuart McGregor, and Nathan Ingold), Taiwan Biobank (Yen-Feng Lin, Yen-Chen Feng, and Hailiang Huang), UCLA Precision Health Biobank (Ruth Johnson, Yi Ding, Alec Chiu, Bogdan Pasaniuc, and Daniel Geschwind), and UK Biobank (Konrad Karczewski and Alicia Martin). The Trøndelag Health Study (HUNT) is a collaboration between HUNT Research Centre (Faculty of Medicine and Health Sciences, NTNU, Norwegian University of Science and Technology), Trøndelag County Council, Central Norway Regional Health Authority, and the Norwegian Institute of Public Health. The genotyping in HUNT was financed by the National Institutes of Health; University of Michigan; the Research Council of Norway; the Liaison Committee for Education, Research and Innovation in Central Norway; and the Joint Research Committee between St Olavs hospital and the Faculty of Medicine and Health Sciences, NTNU. The genetic investigations of the HUNT Study is a collaboration between researchers from the K.G. Jebsen Center for Genetic Epidemiology, NTNU and the University of Michigan Medical School, and the University of Michigan School of Public Health. The K.G. Jebsen Center for Genetic Epidemiology is financed by Stiftelsen Kristian Gerhard Jebsen; Faculty of Medicine and Health Sciences, NTNU, Norway. We want to thank clinicians and other employees at Nord-Trøndelag Hospital Trust, Norway for their support and for contributing to data collection in this research project.

## Author Contributions

Study design: IS, WZ, MJD, BMN, CJW

Bioinformatic analysis: IS, KHW, BNW, WZ, AB, YH, BB

HUNT: IS, BB, AH, MEG, KH

TOPMed replication: YH, CK MVP

replication: JEH, PWFW

Writing: IS, KHW, FS, AB, WH, NRS, CJW

Revision: IS, KHW, BNW, FS, WH, RGW, MJD, SZ, NRS, BMN, CJW

## Declaration of interests

NRS is an advisor for Abbott, Philips, and Shockwave and have received honoraria for speaking from Zoll, Cordis. CJW’s spouse works at Regeneron pharmaceuticals.

## STAR METHODS

### Multi-Ancestry Meta-Analysis

The GBMI ischemic stroke meta-analysis was conducted from genome-wide association results of 16 biobanks using inverse variance weighted meta-analysis. The overall stroke dataset has 1.9% of African/ African American (AFR), 19.6% of East Asian (EAS), 75.8% of European (non-Finnish European: NFE and Finns: FIN), 1.1% of Latino or admixed American (AMR), and 1.6% of South Asian (SAS) ancestry (**Figure 1**). A detailed description of the meta-analysis methods can be found here (Zhou *et al*., 2021). The GBMI stroke phenotype was defined using PheCode 433.21 (Cerebral artery occlusion, with cerebral infarction).

### Polygenic Risk Scores

For the sex-specificity testing, we calculated 3 PRSs using summary statistics from the overall population (N = 1,370,901; 4.3% cases), female only population (N = 601,704; 3.8% cases), and male only population (N = 498,162; 6.1% cases) using PRS-CS (Ge et al., 2019) with a LD reference panel based on combined 1000 Genomes and UK Biobank. The summary statistics used for the PRS calculation excluded the PRS test cohort, HUNT.

### Longitudinal PRS methods in HUNT

HUNT (Krokstad et al., 2013) is a population-based dataset with longitudinal hospital registries linked to the whole genome data (Brumpton et al., 2021). For the PRS prediction in HUNT, we defined IS with ICD9-codes 434 and 436, and ICD10-codes I63 and I64, resulting in 4,256 ischemic stroke cases (285 prevalent, 3,971 incident, 62,375 non-cases). Individuals with no prevalent cardiovascular disease events and full baseline information (non-missing lipid measurements, smoking, anthropometric measures and blood pressure medication information) were included in the survival analysis and the time of event was recorded based on the first appearance of the above-mentioned ICD-codes from both hospital and death registry. The final number of cases in the Cox Proportional Hazards model was 1,796 for males (28,216 non-cases) and 1,858 for females (32,724 non-cases). The survival was modeled with follow-up time as a time-scale with HUNT collection (HUNT2 or HUNT3) as a covariate to count for the possible periodic bias. Individuals that diseased or had a non-ischemic stroke during the follow-up were censored. The PRSs were adjusted with the first ten genetic PCs (for males and females separately for the sex-specific analysis) to remove possible population stratification and the resulting residuals were inverse normalized. All longitudinal analyses were performed using R v4.1.2.

### Replication cohorts

Replication of all seven novel lead variants were requested from two additional GBMI cohorts, Penn Medicine Biobank (PMBB) and Canadian Partnership for Tomorrow’s Health (CanPath). Analyses in these two cohorts were performed using the standard GBMI analysis pipeline (Zhou *et al*., 2021). Furthermore, we received replication results from Million Veteran Program (MVP) and Trans-Omics for Precision Medicine (TOPMed) Program. The MVP (Hunter-Zinck et al., 2020; Klarin et al., 2018) analysis was performed using plink2a (Chang et al., 2015) and with the EHR based stroke phenotype and covariates defined (Klarin *et al*., 2018). Analysis was completed within each HARE-defined ancestry group (Fang et al., 2019). The TOPMed results have been previously published and the analysis details can be found from the original publication (Hu et al., 2021). As TOPMed includes one overlapping biobank with GBMI (BioMe), the GBMI results used to combine discovery and replication excluded the BioMe results.

## Notes

### Competing Interest Statement

NRS is an advisor for Abbott, Philips, and Shockwave and have received honoraria for speaking from Zoll Cordis. The spouse of CJW works at Regeneron pharmaceuticals.

### Funding Statement

Provided as a separate PDF.

### Author Declarations

Participation in the HUNT Study is based on informed consent and the study has been approved by the Data Inspectorate and the Regional Ethics Committee for Medical Research in Norway (REK: 2014/144)

