## Supplementary material for "Multi-ancestry meta-analysis identifies 2 novel loci associated with ischemic stroke and reveals heterogeneity of effects between sexes and ancestries": GBMI Author Banner

Wei Zhou[1,2,3], Masahiro Kanai[1,2,3,4,5], Kuan-Han H Wu[6], Humaira Rasheed[7,8,9], Kristin Tsuo[1,2,3], Jibril B Hirbo[10,11], Ying Wang[1,2,3], Arjun Bhattacharya[12], Huiling Zhao[9], Shinichi Namba[5], Ida Surakka[13], Brooke N Wolford[6,7], Valeria Lo Faro[14,15,16], Esteban A Lopera-Maya[17], Kristi Läll[18], Marie-Julie Favé[19], Sinéad B Chapman[2,3], Juha Karjalainen[1,2,3,20], Mitja Kurki[1,2,3,20], Maasha Mutaamba[1,2,3,20], Juulia Partanen[20], Ben M Brumpton[7,21,22], Sameer Chavan[23], Tzu-Ting Chen[24], Michelle Daya[23], Yi Ding[12,25], Yen-Chen A Feng[26,27], Christopher R Gignoux[23], Sarah E Graham[13], Whitney E Hornsby[13], Nathan Ingold[28,29], Ruth Johnson[12,30], Triin Laisk[18], Kuang Lin[31], Jun Lv[32], Iona Y Millwood[31,33], Priit Palta[18,20], Anita Pandit[34], Michael H Preuss[35], Unnur Thorsteinsdottir[36], Jasmina Uzunovic[19], Matthew Zawistowski[34], Xue Zhong[10,11], Archie Campbell[37], Kristy Crooks[23], Geertruida H de Bock[38], Nicholas J Douville[39,40], Sarah Finer[41], Lars G Fritsche[34], Christopher J Griffiths[41], Yu Guo[42], Karen A Hunt[43], Takahiro Konuma[5,44], Riccardo E Marioni[37], Jansonius Nomdo[14], Snehal Patil[34], Nicholas Rafaels[23], Anne Richmond[45], Jonathan A Shortt[23], Peter Straub[10,11], Ran Tao[11], Brett Vanderwerff[34], Kathleen C Barnes[23], Marike Boezen‡[38], Zhengming Chen[31,33], Chia-Yen Chen[46], Judy Cho[35], George Davey Smith[9,47], Hilary K Finucane[1,2,3], Lude Franke[17], Eric R Gamazon[10,11,48], Andrea Ganna[1,2,20], Tom R Gaunt[9], Tian Ge[27,49], Hailiang Huang[1,2], Jennifer Huffman[50], Jukka T. Koskela[20], Clara Lajonchere[51,52], Matthew H Law[28,29], Liming Li[32], Cecilia M Lindgren[53], Ruth JF Loos[35,54], Stuart MacGregor[28], Koichi Matsuda[55], Catherine M Olsen[28], David J Porteous[37], Jordan A Shavit[56], Harold Snieder[38], Richard C Trembath[57], Judith M Vonk[38], David Whiteman[28], Stephen J Wicks[23], Cisca Wijmenga[17], John Wright[58], Jie Zheng[9], Xiang Zhou[34], Philip Awadalla[19,59], Michael Boehnke[34], Nancy J Cox[10,11], Daniel H Geschwind[51,60,61], Caroline Hayward[45], Kristian Hveem[7,21], Eimear E Kenny[62], Yen-Feng Lin[24,63,64], Reedik Mägi[18], Hilary C Martin[65], Sarah E Medland[28], Yukinori Okada[5,66,67,68,69], Aarno V Palotie[1,2,20], Bogdan Pasaniuc[12,25,51,60,70], Serena Sanna[17,71], Jordan W Smoller[27], Kari Stefansson[36], David A van Heel[43], Robin G Walters[31,33], Sebastian Zöllner[34], Biobank Japan, BioMe, BioVU, Canadian Partnership for Tomorrow's Health/Ontario Health Study, China Kadoorie Biobank Collaborative Group, Colorado Center for Personalized Medicine, deCODE Genetics, Estonian Biobank, FinnGen, Generation Scotland, Genes & Health, LifeLines, Mass General Brigham Biobank, Michigan Genomics Initiative, QIMR Berghofer Biobank, Taiwan Biobank, The HUNT Study, UCLA ATLAS Community Health Initiative, UK Biobank, Alicia R Martin[1,2,3], Cristen J Willer[6,13,72*], Mark J Daly[1,2,3,20*], Benjamin M Neale[1,2,3*]

‡Deceased

*These authors jointly supervised this work

[1]Analytic and Translational Genetics Unit, Department of Medicine, Massachusetts General Hospital, Boston, MA, USA, [2]Stanley Center for Psychiatric Research, Broad Institute of MIT and Harvard, Cambridge, MA, USA, [3]Program in Medical and Population Genetics, Broad Institute of MIT and Harvard, Cambridge, MA, USA, [4]Department of Biomedical Informatics, Harvard Medical School, Boston, MA, USA, [5]Department of Statistical Genetics, Osaka University Graduate School of Medicine, Suita 565-0871, Japan, [6]Department of Computational Medicine and Bioinformatics, University of Michigan, Ann Arbor, MI, USA, [7]K.G. Jebsen Center for Genetic Epidemiology, Department of Public Health and Nursing, NTNU, Norwegian University of Science and Technology, Trondheim, Norway, [8]Division of Medicine and Laboratory Sciences, University of Oslo, Norway, [9]MRC Integrative Epidemiology Unit (IEU), Bristol Medical School, University of Bristol, Bristol, UK, [10]Department of Medicine, Division of Genetic Medicine, Vanderbilt University Medical Center, Nashville, TN, USA, [11]Vanderbilt Genetics Institute, Vanderbilt University

Medical Center, Nashville, TN, USA, [12]Department of Pathology and Laboratory Medicine, David Geffen School of Medicine, University of California, Los Angeles, Los Angeles, CA, USA, [13]Department of Internal Medicine, Division of Cardiology, University of Michigan, Ann Arbor, MI, USA, [14]University of Groningen, UMCG, Department of Ophthalmology, Groningen, the Netherlands, [15]Department of Clinical Genetics, Amsterdam University Medical Center (AMC), Amsterdam, the Netherlands, [16]Department of Immunology, Genetics and Pathology, Science for Life Laboratory, Uppsala University, Uppsala, Sweden, [17]University of Groningen, UMCG, Department of Genetics, Groningen, the Netherlands, [18]Estonian Genome Centre, Institute of Genomics, University of Tartu, Tartu, Estonia, [19]Ontario Institute for Cancer Research, Toronto, ON, Canada, [20]Institute for Molecular Medicine Finland, University of Helsinki, Helsinki, Finland, [21]HUNT Research Centre, Department of Public Health and Nursing, NTNU, Norwegian University of Science and Technology, Levanger, Norway, [22]Clinic of Medicine, St. Olavs Hospital, Trondheim University Hospital, Trondheim, Norway, [23]University of Colorado - Anschutz Medical Campus, Aurora, CO, USA, [24]Center for Neuropsychiatric Research, National Health Research Institutes, Miaoli, Taiwan, [25]Bioinformatics Interdepartmental Program, University of California, Los Angeles, Los Angeles, CA, USA, [26]Division of Biostatistics, Institute of Epidemiology and Preventive Medicine, College of Public Health, National Taiwan University, Taiwan, [27]Psychiatric and Neurodevelopmental Genetics Unit, Center for Genomic Medicine, Massachusetts General Hospital, Boston, MA, USA, [28]QIMR Berghofer Medical Research Institute, Brisbane, Australia, [29]Faculty of Health, School of Biomedical Sciences, Queensland University of Technology, Australia, [30]Department of Computer Science, University of California, Los Angeles, Los Angeles, CA, USA, [31]Nuffield Department of Population Health, University of Oxford, Oxford, UK, [32]Department of Epidemiology and Biostatistics, School of Public Health, Peking University Health Science Center, Beijing, China, [33]MRC Population Health Research Unit, University of Oxford, Oxford, UK, [34]Department of Biostatistics and Center for Statistical Genetics, University of Michigan, Ann Arbor, MI, USA, [35]The Charles Bronfman Institute for Personalized Medicine, Icahn School of Medicine at Mount Sinai, New York, NY, USA, [36]deCODE Genetics/Amgen inc., 101, Reykjavik, Iceland, [37]Centre for Genomic and Experimental Medicine, Institute of Genetics and Cancer, University of Edinburgh, Edinburgh, UK, [38]University of Groningen, UMCG, Department of Epidemiology, Groningen, the Netherlands, [39]Department of Anesthesiology, Michigan Medicine, Ann Arbor, MI, USA, [40]Institute of Healthcare Policy & Innovation, University of Michigan, Ann Arbor, MI, USA, [41]Wolfson Institute of Population Health, Queen Mary University of London, London, UK, [42]Chinese Academy of Medical Sciences, Beijing, China, [43]Blizard Institute, Queen Mary University of London, London, UK, [44]Central Pharmaceutical Research Institute, JAPAN TOBACCO INC., Takatsuki 569-1125, Japan, [45]Medical Research Council Human Genetics Unit, Institute of Genetics and Cancer, University of Edinburgh, Edinburgh, UK, [46]Biogen, Cambridge, MA, USA, [47]NIHR Bristol Biomedical Research Centre, Bristol, UK, [48]MRC Epidemiology Unit, University of Cambridge, Cambridge, UK, [49]Center for Precision Psychiatry, Massachusetts General Hospital, Boston, MA, USA, [50]Centre for Population Genomics, VA Boston Healthcare System, Boston, MA, USA, [51]Institute of Precision Health, University of California, Los Angeles, Los Angeles, CA, USA, [52]Program in Neurogenetics, Department of Neurology, David Geffen School of Medicine, University of California, Los Angeles, Los Angeles, CA, USA, [53]Big Data Institute, Li Ka Shing Centre for Health Information and Discovery, University of Oxford, Oxford, UK, [54]Novo Nordisk Foundation Center for Basic Metabolic Research, Faculty of Medicine and Health Sciences, University of Copenhagen, Copenhagen, Denmark, [55]Department of Computational Biology and Medical Sciences, Graduate school of Frontier Sciences, The University of Tokyo, Tokyo, Japan,

[56]University of Michigan, Department of Pediatrics, Ann Arbor MI 48109, [57]School of Basic and Medical Biosciences, Faculty of Life Sciences and Medicine, King's College London, London, UK, [58]Bradford Institute for Health Research, Bradford Teaching Hospitals National Health Service (NHS) Foundation Trust, Bradford, UK, [59]Department of Molecular Genetics, University of Toronto, Toronto, ON, Canada, [60]Department of Human Genetics, David Geffen School of Medicine, University of California, Los Angeles, Los Angeles, CA, USA, [61]Department of Neurology, David Geffen School of Medicine, University of California, Los Angeles, Los Angeles, CA, USA, [62]Institute for Genomic Health, Icahn School of Medicine at Mount Sinai, New York, NY, USA, [63]Department of Public Health & Medical Humanities, School of Medicine, National Yang Ming Chiao Tung University, Taipei, Taiwan, [64]Institute of Behavioral Medicine, College of Medicine, National Cheng Kung University, Tainan, Taiwan, [65]Medical and Population Genomics, Wellcome Sanger Institute, Hinxton, UK, [66]Center for Infectious Disease Education and Research (CiDER), Osaka University, Suita 565-0871, Japan, [67]Laboratory of Statistical Immunology, Immunology Frontier Research Center (WPI-IFReC), Osaka University, Suita 565-0871, Japan, [68]Laboratory for Systems Genetics, RIKEN Center for Integrative Medical Sciences, Yokohama, Japan, [69]Integrated Frontier Research for Medical Science Division, Institute for Open and Transdisciplinary Research Initiatives, Osaka University, Suita 565-0871, Japan, [70]Department of Computational Medicine, David Geffen School of Medicine, University of California, Los Angeles, Los Angeles, CA, USA, [71]Institute for Genetics and Biomedical Research (IRGB), National Research Council (CNR), Cagliari, Italy, [72]Department of Human Genetics, University of Michigan, Ann Arbor, MI, USA
