## Supplementary material for "Multi-ancestry meta-analysis identifies 2 novel loci associated with ischemic stroke and reveals heterogeneity of effects between sexes and ancestries": GBMI Author Acknowledgements

### Acknowledgments

#### Biobank Japan Project
The BioBank Japan Project was supported by the Tailor-Made Medical Treatment program of the Ministry of Education, Culture, Sports, Science, and Technology (MEXT), the Japan Agency for Medical Research and Development (AMED).
S.N. was supported by Takeda Science Foundation.
Y.O. was supported by JSPS KAKENHI (19H01021, 20K21834), and AMED (JP21km0405211, JP21ek0109413, JP21ek0410075, JP21gm4010006, and JP21km0405217), JST Moonshot R&D (JPMJMS2021, JPMJMS2024), Takeda Science Foundation, and Bioinformatics Initiative of Osaka University Graduate School of Medicine, Osaka University.

#### BioMe - The Mount Sinai BioMe Biobank
The Mount Sinai BioMe Biobank has been supported by The Andrea and Charles Bronfman Philanthropies and in part by Federal funds from the NHLBI and NHGRI (U01HG00638001; U01HG007417; X01HL134588). We thank all participants in the Mount Sinai Biobank. We also thank all our recruiters who have assisted and continue to assist in data collection and management and are grateful for the computational resources and staff expertise provided by Scientific Computing at the Icahn School of Medicine at Mount Sinai.

#### BioVU
The BioVU projects at Vanderbilt University Medical Center are supported by numerous sources: institutional funding, private agencies, and federal grants. These include the NIH-funded Shared Instrumentation Grant S10OD017985 and S10RR025141; CTSA grants UL1TR002243, UL1TR000445, and UL1RR024975 from the National Center for Advancing Translational Sciences. Its contents are solely the responsibility of the authors and do not necessarily represent official views of the National Center for Advancing Translational Sciences or the National Institutes of Health. Genomic data are also supported by investigator-led projects that include U01HG004798, R01NS032830, RC2GM092618, P50GM115305, U01HG006378, U19HL065962, R01HD074711; and additional funding sources listed at https://victr. vumc.org/biovu-funding/.

#### Canadian Partnership for Tomorrow's Health/Ontario Health Study
This research was supported by the Canadian Institute of Health Research (CIHR) grant #EC3-144623

#### Colorado Center for Personalized Medicine (CCPM)
The Colorado Center for Personalized Medicine (CCPM) would like to thank Richard Zane, Steve Hess, Sarah White, Emily Hearst, Emily Roberts and the entire Health Data Compass team. CCPM was developed with support from UCHealth, Children's Hospital Colorado, CU Medicine, CU Department of Medicine and CU School of Medicine.

**China Kadoorie Biobank collaborative group**
International Steering Committee: Junshi Chen, Zhengming Chen (PI), Robert Clarke, Rory Collins, Yu Guo, Liming Li (PI), Jun Lv, Richard Peto, Robin Walters, Chen Wang.

*International Co-ordinating Centre, Oxford:* Daniel Avery, Fiona Bragg, Derrick Bennett, Ruth Boxall, Ka Hung Chan, Yumei Chang, Yiping Chen, Zhengming Chen, Johnathan Clarke; Robert Clarke, Huaidong Du, Zammy Fairhurst-Hunter, Hannah Fry, Simon Gilbert, Alex Hacker, Parisa Hariri, Mike Hill, Michael Holmes, Pek Kei Im, Andri Iona, Maria Kakkoura, Christiana Kartsonaki, Rene Kerosi, Kuang Lin, Mohsen Mazidi, Iona Millwood, Qunhua Nie, Alfred Pozarickij, Paul Ryder, Sam Sansome, Dan Schmidt, Paul Sherliker, Rajani Sohoni, Becky Stevens, Iain Turnbull, Robin Walters, Lin Wang, Neil Wright, Ling Yang, Xiaoming Yang, Pang Yao.

National Co-ordinating Centre, Beijing: Yu Guo, Xiao Han, Can Hou, Chun Li, Chao Liu, Jun Lv, Pei Pei, Canqing Yu.

*Regional Coordinating Centres:*

Guangxi Provincial CDC: Naying Chen, Duo Liu, Zhenzhu Tang. **Liuzhou CDC:** Ningyu Chen, Qilian Jiang, Jian Lan, Mingqiang Li, Yun Liu, Fanwen Meng, Jinhuai Meng, Rong Pan, Yulu Qin, Ping Wang, Sisi Wang, Liuping Wei, Liyuan Zhou. **Gansu Provincial CDC:** Caixia Dong, Pengfei Ge, Xiaolan Ren. **Maiji CDC:** Zhongxiao Li, Enke Mao, Tao Wang, Hui Zhang, Xi Zhang. **Hainan Provincial CDC:** Jinyan Chen, Ximin Hu, Xiaohuan Wang. **Meilan CDC:** Zhendong Guo, Huimei Li, Yilei Li, Min Weng, Shukuan Wu. **Heilongjiang Provincial CDC:** Shichun Yan, Mingyuan Zou, Xue Zhou. **Nangang CDC:** Ziyan Guo, Quan Kang, Yanjie Li, Bo Yu, Qinai Xu. **Henan Provincial CDC:** Liang Chang, Lei Fan, Shixian Feng, Ding Zhang, Gang Zhou. **Huixian CDC:** Yulian Gao, Tianyou He, Pan He, Chen Hu, Huarong Sun, Xukui Zhang. **Hunan Provincial CDC:** Biyun Chen, Zhongxi Fu, Yuelong Huang, Huilin Liu, Qiaohua Xu, Li Yin. **Liuyang CDC:** Huajun Long, Xin Xu, Hao Zhang, Libo Zhang. **Jiangsu Provincial CDC:** Jian Su, Ran Tao, Ming Wu, Jie Yang, Jinyi Zhou, Yonglin Zhou. **Suzhou CDC:** Yihe Hu, Yujie Hua, Jianrong Jin Fang Liu, Jingchao Liu, Yan Lu, Liangcai Ma, Aiyu Tang, Jun Zhang. **Qingdao CDC:** Liang Cheng, Ranran Du, Ruqin Gao, Feifei Li, Shanpeng Li, Yongmei Liu, Feng Ning, Zengchang Pang, Xiaohui Sun, Xiaocao Tian, Shaojie Wang, Yaoming Zhai, Hua Zhang. **Licang CDC:** Wei Hou, Silu Lv, Junzheng Wang. **Sichuan Provincial CDC:** Xiaofang Chen, Xianping Wu, Ningmei Zhang, Weiwei Zhou. **Pengzhou CDC:** Xiaofang Chen, Jianguo Li, Jiaqiu Liu, Guojin Luo, Qiang Sun, Xunfu Zhong. **Zhejiang Provincial CDC:** Weiwei Gong, Ruying Hu, Hao Wang,Meng Wan, Min Yu. **Tongxiang CDC:** Lingli Chen, Qijun Gu, Dongxia Pan, Chunmei Wang, Kaixu Xie, Xiaoyi Zhang.

CKB Acknowledgements and Funding:

China Kadoorie Biobank gratefully acknowledges the participants, project staff, and the China National Centre for Disease Control and Prevention (CDC) and its regional offices. China's National Health Insurance provides electronic linkage to all hospital treatment. Funding sources: Baseline survey and first re-survey – Kadoorie Charitable Foundation, Hong Kong; long-term follow-up – UK Wellcome Trust (212946/Z/18/Z, 202922/Z/16/Z, 104085/Z/14/Z,

088158/Z/09/Z), National Natural Science Foundation of China (91843302), National Key Research and Development Program of China (2016YFC 0900500, 0900501, 0900504, 1303904); DNA extraction and genotyping – GlaxoSmithKline, UK Medical Research Council (MC-PC-13049, MC-PC-14135); core funding for the project to the Clinical Trial Service Unit and Epidemiological Studies Unit at Oxford University – British Heart Foundation (CH/1996001/9454), UK Medical Research Council (MC-UU-00017/1, MC-UU-12026/2, MC_U137686851), Cancer Research UK (C16077/A29186, C500/A16896).

**deCODE Genetics**
We thank participants in deCODE cardiovascular and obesity studies and collaborators for their cooperation.

**Estonian Biobank**
This research was supported by the European Union through Horizon 2020 research and innovation programme under grant no 810645 and through the European Regional Development Fund project no. MOBEC008, by the Estonian Research Council grant PUT (PRG1291, PRG687 and PRG184) and by the European Union through the European Regional Development Fund project no. MOBERA21 (ERA-CVD project DETECT ARRHYTHMIAS, GA no JTC2018-009), Project No. 2014-2020.4.01.15-0012 and Project No. 2014-2020.4.01.16-0125.
We would like to acknowledge Dr. Tõnu Esko; Dr. Lili Milani; Dr. Reedik Mägi, Dr. Mari Nelis and Dr. Andres Metspalu, all from the Institute of Genomics, University of Tartu, Tartu, Estonia.

**FinnGen**
The FinnGen project is funded by two grants from Business Finland (HUS 4685/31/2016 and UH 4386/31/2016) and the following industry partners: AbbVie Inc., AstraZeneca UK Ltd, Biogen MA Inc., Bristol Myers Squibb (and Celgene Corporation & Celgene International II Sàrl), Genentech Inc., Merck Sharp & Dohme Corp, Pfizer Inc., GlaxoSmithKline Intellectual Property Development Ltd., Sanofi US Services Inc., Maze Therapeutics Inc., Janssen Biotech Inc, and Novartis AG. Following biobanks are acknowledged for delivering biobank samples to FinnGen: Auria Biobank (www.auria.fi/biopankki), THL Biobank (www.thl.fi/biobank), Helsinki Biobank (www.helsinginbiopankki.fi), Biobank Borealis of Northern Finland (https://www.ppshp.fi/Tutkimus-ja-opetus/Biopankki/Pages/Biobank-Borealis-briefly-in-English.aspx), Finnish Clinical Biobank Tampere (www.tays.fi/en-US/Research_and_development/Finnish_Clinical_Biobank_Tampere), Biobank of Eastern Finland (www.ita-suomenbiopankki.fi/en), Central Finland Biobank (www.ksshp.fi/fi-FI/Potilaalle/Biopankki), Finnish Red Cross Blood Service Biobank (www.veripalvelu.fi/verenluovutus/biopankkitoiminta) and Terveystalo Biobank (www.terveystalo.com/fi/Yritystietoa/Terveystalo-Biopankki/Biopankki/). All Finnish Biobanks are members of BBMRI.fi infrastructure (www.bbmri.fi). Finnish Biobank Cooperative -FINBB (https://finbb.fi/) is the coordinator of BBMRI-ERIC operations in Finland. The Finnish biobank data can be accessed through the Fingenious® services (https://site.fingenious.fi/en/) managed by FINBB.

**Generation Scotland**
Generation Scotland received core support from the Chief Scientist Office of the Scottish Government Health Directorates [CZD/16/6] and the Scottish Funding Council [HR03006] and is currently supported by the Wellcome Trust [216767/Z/19/Z]. Genotyping of the GS:SFHS samples was carried out by the Genetics Core Laboratory at the Edinburgh Clinical Research Facility, University of Edinburgh, Scotland and was funded by the Medical Research Council UK and the Wellcome Trust (Wellcome Trust Strategic Award "STratifying Resilience and Depression Longitudinally" (STRADL) Reference 104036/Z/14/Z)."

**Genes and Health**
Genes & Health is/has recently been core-funded by Wellcome (WT102627, WT210561), the Medical Research Council (UK) (M009017), Higher Education Funding Council for England Catalyst, Barts Charity (845/1796), Health Data Research UK (for London substantive site), and research delivery support from the NHS National Institute for Health Research Clinical Research Network (North Thames). We thank Social Action for Health, Centre of The Cell, members of our Community Advisory Group, and staff who have recruited and collected data from volunteers. We thank the NIHR National Biosample Centre (UK Biocentre), the Social Genetic & Developmental Psychiatry Centre (King's College London), Wellcome Sanger Institute, and Broad Institute for sample processing, genotyping, sequencing and variant annotation. We thank: Barts Health NHS Trust, NHS Clinical Commissioning Groups (City and Hackney, Waltham Forest, Tower Hamlets, Newham, Redbridge, Havering, Barking and Dagenham), East London NHS Foundation Trust, Bradford Teaching Hospitals NHS Foundation

Trust, Public Health England (especially David Wyllie), Discovery Data Service/Endeavour Health Charitable Trust (especially David Stables) - for GDPR-compliant data sharing backed by individual written informed consent. Most of all we thank all of the volunteers participating in Genes & Health.

Genes & Health Research Team (in alphabetical order by surname): Shaheen Akhtar, Mohammad Anwar, Elena Arciero, Samina Ashraf, Gerome Breen, Raymond Chung, Charles J Curtis, Maharun Chowdhury, Grainne Colligan, Panos Deloukas, Ceri Durham, Sarah Finer, Chris Griffiths, Qin Qin Huang, Matt Hurles, Karen A Hunt, Shapna Hussain, Kamrul Islam, Ahsan Khan, Amara Khan, Cath Lavery, Sang Hyuck Lee, Robin Lerner, Daniel MacArthur, Bev MacLaughlin, Hilary Martin, Dan Mason, Shefa Miah, Bill Newman, Nishat Safa, Farah Tahmasebi, Richard C Trembath, Bhavi Trivedi, David A van Heel, John Wright.

**The HUNT Study**

A special thanks to all the HUNT participants for donating their time, samples and information to help others.

The Trøndelag Health Study (HUNT) is a collaboration between HUNT Research Centre (Faculty of Medicine and Health Sciences, NTNU, Norwegian University of Science and Technology), Trøndelag County Council, Central Norway Regional Health Authority, and the Norwegian Institute of Public Health. The genotyping in HUNT was financed by the National Institutes of Health; University of Michigan; the Research Council of Norway; the Liaison Committee for Education, Research and Innovation in Central Norway; and the Joint Research Committee between St Olavs hospital and the Faculty of Medicine and Health Sciences, NTNU. The genetic investigations of the HUNT Study, is a collaboration between researchers from the K.G. Jebsen Center for Genetic Epidemiology, NTNU and the University of Michigan Medical School and the University of Michigan School of Public Health. The K.G. Jebsen Center for Genetic Epidemiology is financed by Stiftelsen Kristian Gerhard Jebsen; Faculty of Medicine and Health Sciences, NTNU, Norway.

We want to thank clinicians and other employees at Nord-Trøndelag Hospital Trust for their support and for contributing to data collection in this research project.

We also acknowledge; **HUNT-MI Leadership:** Kristian Hveem, Cristen Willer, Oddgeir Lingaas Holmen, Mike Boehnke, Goncalo Abecasis, Bjorn Olav Åsvold, Ben Brumpton; **Scientific Advisory Committee:** Ele Zeggini, Mark Daly, Bjørn Pasternak; **HUNT Research Centre:** Jørn Søberg Fenstad, Anne Jorunn Vikdal, Marit Næss; **HUNT Cloud:** Oddgeir Lingaas Holmen, Sandor Zeestraten, Tom Erik Røberg; **Data applications and registry linkages:** Maiken E. Gabrielsen, Anne Heidi Skogholt; **Low-pass whole sequencing genome bioinformatics and statistical analysis:** He Zhang, Hyun Min Kang, Jin Chen; **Array genotyping:** Sten Even Erlandsen, Vidar Beisvåg; **GWAS bioinformatics, QC, imputation and statistical analysis:** Wei Zhou, Jonas Nielsen, Lars Fritsche, Hyun Min Kang, Oddgeir Holmen, Ben Brumpton,

Laurent Thomas; **CNV calling:** Ellen Schmidt, Ryan Mills; **Statistical methods development for analyzing HUNT data:** Wei Zhou, Shawn Lee

**Funding:**
The K. G. Jebsen Centre for Genetic Epidemiology is financed by Stiftelsen Kristian Gerhard Jebsen. The genotyping in HUNT was financed by the National Institutes of Health; University of Michigan; the Research Council of Norway; Stiftelsen Kristian Gerhard Jebsen; the Liaison Committee for Education, Research and Innovation in Central Norway; and the Joint Research Committee between St Olav's hospital and the Faculty of Medicine and Health Sciences, NTNU.

**LifeLines**
The Lifelines Biobank initiative has been made possible by funding from the Dutch Ministry of Health, Welfare and Sport, the Dutch Ministry of Economic Affairs, the University Medical Center Groningen (UMCG the Netherlands), University of Groningen and the Northern Provinces of the Netherlands. The generation and management of GWAS genotype data for the Lifelines Cohort Study is supported by the UMCG Genetics Lifelines Initiative (UGLI). UGLI is partly supported by a Spinoza Grant from NWO, awarded to Cisca Wijmenga.

The authors wish to acknowledge the services of the Lifelines Cohort Study, the contributing research centers delivering data to Lifelines, and all the study participants.

*UMCG Genetics Lifelines Initiative (UGLI) team:* Raul Aguirre-Gamboa (1), Patrick Deelen (1), Lude Franke (1), Jan A Kuivenhoven (2), Esteban A Lopera Maya (1), Ilja M Nolte (3), Serena Sanna (1), Harold Snieder (3), Morris A Swertz (1), Peter M. Visscher (3,4), Judith M Vonk (3), Cisca Wijmenga (1)

(1) *Department of Genetics, University of Groningen, University Medical Center Groningen, The Netherlands*

(2) *Department of Pediatrics, University of Groningen, University Medical Center Groningen, The Netherlands*

(3) *Department of Epidemiology, University of Groningen, University Medical Center Groningen, The Netherlands*

(4) *Institute for Molecular Bioscience, The University of Queensland, Brisbane, Queensland, Australia.*

**Mass General Brigham (MGB) Biobank**
Samples, genomic data, and health information were obtained from the Mass General Brigham Biobank, a biorepository of consented patients samples at Mass General Brigham (parent organization of Massachusetts General Hospital and Brigham and Women's Hospital). We are grateful to all of the participants and clinical and research teams who made this work possible.

Support for genotyping was provided through MGB Personalized Medicine.

MGB Biobank Leadership: Elizabeth W. Karlson, MD; Shawn N. Murphy, MD, PhD; Susan A Slaugenhaupt, PhD; Jordan W. Smoller, MD, ScD; Scott T. Weiss, MD, MSc

**Michigan Genomics Initiative**
The authors acknowledge the Michigan Genomics Initiative participants, Precision Health at the University of Michigan, the University of Michigan Medical School Central Biorepository, and the University of Michigan Advanced Genomics Core for providing data and specimen storage, management, processing, and distribution services, and the Center for Statistical Genetics in the Department of Biostatistics at the School of Public Health for genotype data curation, imputation, and management in support of the research reported in this publication.

**QIMR Berghofer Biobank**
This work is supported by a project grant (APP1063061) and a Program grant (APP1073898) from the Australian National Health and Medical Research Council (NHMRC). SM is supported by a Research Fellowship from the NHMRC (Aust). NI received scholarship support from the University of Queensland and QIMR Berghofer Medical Research Institute. We thank research staff (David Whiteman, Catherine Olsen, Rachel Neale) and participants from the Australian QSKIN study (https://www.qimrberghofer.edu.au/study/qskin/).
SEM is supported in part by APP1172917 from the Australian National Health and Medical Research Council (NHMRC).

**Taiwan Biobank**
This research has been conducted using the Taiwan Biobank resource. We thank all the participants and investigators of the Taiwan Biobank. We thank the National Center for Genome Medicine of Taiwan for the technical support in genotyping. We thank the National Core Facility for Biopharmaceuticals (NCFB, MOST 106-2319-B-492-002) and National Center for High-performance Computing (NCHC) of National Applied Research Laboratories (NARLabs) of Taiwan for providing computational and storage resources.

**UCLA ATLAS Community Health Initiative (UCLA)**
We gratefully acknowledge the resources provided by the Institute for Precision Health (IPH) and participating UCLA ATLAS Community Health Initiative patients. The UCLA ATLAS Community Health Initiative in collaboration with UCLA ATLAS Precision Health Biobank, is a program of IPH, which directs and supports the biobanking and genotyping of biospecimen samples from participating UCLA patients in collaboration with the David Geffen School of Medicine, UCLA CTSI and UCLA Health. Members of the UCLA ATLAS Community Health Initiative include Ruth Johnson, Yi Ding, Vidhya Venkateswaran, Arjun Bhattacharya, Alec Chiu, Tommer Schwarz, Malika Freund, Lingyu Zhan, Kathryn S. Burch, Christa Caggiano, Brian Hill, Nadav Rakocz, Brunilda Balliu, Jae Hoon Sul, Noah Zaitlen, Valerie A. Arboleda, Eran Halperin, Sriram Sankararaman, Manish J. Butte, Clara Lajonchere, Daniel H. Geschwind, and Bogdan Pasaniuc, on behalf of theUCLA Precision Health Data Discovery Repository Working Group

and UCLA Precision Health ATLAS Working Group.

**UK Biobank**
Access to data from the UK BioBank was obtained through Application #31063
PI: Ben Neale, Claire Churchhouse
Overview: “Methodological extensions to estimate genetic heritability and shared risk factors for phenotypes of the UK Biobank”.
Website for Pan-UKBB results can be found: https://pan.ukbb.broadinstitute.org/

**Other**
G.D.S.; T.R.G.; and J.Z. are supported by a grant from the Medical Research Council for the Integrative Epidemiology Unit at the University of Bristol MC_UU_00011/1 & 4.
J.Z. is supported by the Academy of Medical Sciences (AMS) Springboard Award, the Wellcome Trust, the Government Department of Business, Energy and Industrial Strategy (BEIS), the British Heart Foundation and Diabetes UK (SBF006\1117). J.Z. is funded by the Vice-Chancellor Fellowship from the University of Bristol. W.Z. was supported by the National Human Genome Research Institute of the National Institutes of Health under award number T32HG010464.

**ICDA**
The authors would like to acknowledge the organizing committee of the International Common Disease Alliance for intellectual contributions on the set up of the GBMI as a nascent activity to the larger effort. We also thank them for the use of their slack platform.
Website for ICDA can be found here: https://www.icda.bio/

**The Hail Team and Data Management at the Stanley Center for Psychiatric Research**
Hail is an open-source Python library that simplifies genomic data analysis in the cloud. It provides powerful, easy-to-use data science tools that can be used to interrogate biobank-scale genomic data and was used in the analysis of the data for this paper. We would especially like to thank Daniel King from the Hail team and Sam Bryant from the Stanley Center Data Management team for helping with the Google bucket set up and data sharing.
Website for Hail can be found here: https://hail.is/
