## Supplementary Information for "Multi-ancestry meta-analysis identifies 2 novel loci associated with ischemic stroke and reveals heterogeneity of effects between sexes and ancestries"

**Supplementary Note**

**NHLBI Trans-Omics for Precision Medicine (TOPMed) Stroke Working Group Acknowledgements**

***Members:***

Christopher D. Anderson, Tim Assimes, Paul Auer, Alexa Beiser, Joshua Bis, Cara Carty, Kei Hang Katie Chan, Jaeyoon Chung, John Cole, Adolfo Correa, Nicole Davis Armstrong, Ron Do, Zhaohui Du, Nauder Faraday, Myriam Fornage, Jeff Haessler, Scott Heemann, Yao Hu, Rebecca Jackson, Thomas Jaworek, Michelle Kim, Charles Kooperberg, Christina Kourkoulis, Leslie Lange, Simin Liu, Will Longstreth, Jessica Lundin, Tracy Madsen, Sandro Marini, Julie Mikulla, Braxton D. Mitchell, Paul Nyquist, James Perry, Ulrike Peters, Gloria Quach, Vasan S. Ramachandran, Alex Reiner, Stephen Rich, Jonathan Rosand, Jerome Rotter, Muralidharan Sargurupremrajm Chloé Sarnowski, Jenny Schoenberg, Sudha Seshadri, Brian Silver, Sylvia Smoller, David Tirschwell, Dhananjay Vaidya, Joanna von Berg, Huichun Xu, Ying Zhou

***Acknowledgements***

We gratefully acknowledge the studies and participants who provided biological samples and data for TOPMed.

ARIC: The authors thank the staff and participants of the ARIC study for their important contributions.

FHS: We acknowledge the dedication of the FHS study participants without whom this research would not be possible.

JHS: The authors wish to thank the staffs and participants of the JHS.

WHI: The authors thank the WHI investigators and staff for their dedication, and the study participants for making the program possible. A full listing of WHI investigators can be found at: <http://www.whi.org/researchers/Documents%20%20Write%20a%20Paper/WHI%20Investigator%20Long%20List.pdf>.

**Million Veteran Program (MVP) Acknowledgements**

***MVP Executive Committee***

- Co-Chair: J. Michael Gaziano, M.D., M.P.H.

VA Boston Healthcare System, 150 S. Huntington Avenue, Boston, MA 02130

- Co-Chair: Sumitra Muralidhar, Ph.D.

US Department of Veterans Affairs, 810 Vermont Avenue NW, Washington, DC 20420

- Rachel Ramoni, D.M.D., Sc.D., Chief VA Research and Development Officer

US Department of Veterans Affairs, 810 Vermont Avenue NW, Washington, DC 20420

- Jean Beckham, Ph.D.

Durham VA Medical Center, 508 Fulton Street, Durham, NC 27705

- Kyong-Mi Chang, M.D.

Philadelphia VA Medical Center, 3900 Woodland Avenue, Philadelphia, PA 19104

- Philip S. Tsao, Ph.D.

VA Palo Alto Health Care System, 3801 Miranda Avenue, Palo Alto, CA 94304

- James Breeling, M.D., Ex-Officio

US Department of Veterans Affairs, 810 Vermont Avenue NW, Washington, DC 20420

- Grant Huang, Ph.D., Ex-Officio

US Department of Veterans Affairs, 810 Vermont Avenue NW, Washington, DC 20420

- Juan P. Casas, M.D., Ph.D., Ex-Officio

VA Boston Healthcare System, 150 S. Huntington Avenue, Boston, MA 02130

***MVP Program Office***

- Sumitra Muralidhar, Ph.D.

US Department of Veterans Affairs, 810 Vermont Avenue NW, Washington, DC 20420

- Jennifer Moser, Ph.D.

US Department of Veterans Affairs, 810 Vermont Avenue NW, Washington, DC 20420

***MVP Recruitment/Enrollment***

- MVP Cohort Management Director/Recruitment/Enrollment Director, Boston – Stacey B. Whitbourne, Ph.D.; Jessica V. Brewer, M.P.H.

VA Boston Healthcare System, 150 S. Huntington Avenue, Boston, MA 02130

- VA Central Biorepository, Boston – Mary T. Brophy M.D., M.P.H.; Donald E. Humphries, Ph.D.; Luis E. Selva, Ph.D.

VA Boston Healthcare System, 150 S. Huntington Avenue, Boston, MA 02130

- MVP Informatics, Boston – Nhan Do, M.D.; Shahpoor (Alex) Shayan, M.S.

VA Boston Healthcare System, 150 S. Huntington Avenue, Boston, MA 02130

- MVP Data Operations/Analytics, Boston – Kelly Cho, M.P.H., Ph.D.

VA Boston Healthcare System, 150 S. Huntington Avenue, Boston, MA 02130

- Director of Regulatory Affairs – Lori Churby, B.S.

VA Palo Alto Health Care System, 3801 Miranda Avenue, Palo Alto, CA 94304

- MVP Coordinating Centers
  - Cooperative Studies Program Clinical Research Pharmacy Coordinating Center, Albuquerque – Todd Connor, Pharm.D.; Dean P. Argyres, B.S., M.S.

New Mexico VA Health Care System, 1501 San Pedro Drive SE, Albuquerque, NM 87108

- Genomics Coordinating Center, Palo Alto – Philip S. Tsao, Ph.D.

VA Palo Alto Health Care System, 3801 Miranda Avenue, Palo Alto, CA 94304

- MVP Boston Coordinating Center, Boston - J. Michael Gaziano, M.D., M.P.H.

VA Boston Healthcare System, 150 S. Huntington Avenue, Boston, MA 02130

- MVP Information Center, Canandaigua – Brady Stephens, M.S.

Canandaigua VA Medical Center, 400 Fort Hill Avenue, Canandaigua, NY 14424

***MVP Science***

Saiju Pyarajan Ph.D.

VA Boston Healthcare System, 150 S. Huntington Avenue, Boston, MA 02130

Philip S. Tsao, Ph.D.

VA Palo Alto Health Care System, 3801 Miranda Avenue, Palo Alto, CA 94304

- Data Core - Kelly Cho, M.P.H, Ph.D.

VA Boston Healthcare System, 150 S. Huntington Avenue, Boston, MA 02130

- VA Informatics and Computing Infrastructure (VINCI) – Scott L. DuVall, Ph.D.

VA Salt Lake City Health Care System, 500 Foothill Drive, Salt Lake City, UT 84148

- Data and Computational Sciences – Saiju Pyarajan, Ph.D.

VA Boston Healthcare System, 150 S. Huntington Avenue, Boston, MA 02130

- Statistical Genetics – Elizabeth Hauser, Ph.D.

Durham VA Medical Center, 508 Fulton Street, Durham, NC 27705

Yan Sun, Ph.D.

Atlanta VA Medical Center, 1670 Clairmont Road, Decatur, GA 30033

Hongyu Zhao, Ph.D.

West Haven VA Medical Center, 950 Campbell Avenue, West Haven, CT 06516

***Current MVP Local Site Investigators***

- Atlanta VA Medical Center (Peter Wilson, M.D.)

1670 Clairmont Road, Decatur, GA 30033

- Bay Pines VA Healthcare System (Rachel McArdle, Ph.D.)

10,000 Bay Pines Blvd Bay Pines, FL 33744

- Birmingham VA Medical Center (Louis Dellitalia, M.D.)

700 S. 19th Street, Birmingham AL 35233

- Central Western Massachusetts Healthcare System (Kristin Mattocks, Ph.D., M.P.H.)

421 North Main Street, Leeds, MA 01053

- Cincinnati VA Medical Center (John Harley, M.D., Ph.D.)

3200 Vine Street, Cincinnati, OH 45220

- Clement J. Zablocki VA Medical Center (Jeffrey Whittle, M.D., M.P.H.)

5000 West National Avenue, Milwaukee, WI 53295

- VA Northeast Ohio Healthcare System (Frank Jacono, M.D.)

10701 East Boulevard, Cleveland, OH 44106

- Durham VA Medical Center (Jean Beckham, Ph.D.)

508 Fulton Street, Durham, NC 27705

- Edith Nourse Rogers Memorial Veterans Hospital (John Wells., Ph.D.)

200 Springs Road, Bedford, MA 01730

- Edward Hines, Jr. VA Medical Center (Salvador Gutierrez, M.D.)

5000 South 5th Avenue, Hines, IL 60141

- Veterans Health Care System of the Ozarks (Kathrina Alexander, M.D.)

1100 North College Avenue, Fayetteville, AR 72703

- Fargo VA Health Care System (Kimberly Hammer, Ph.D.)

2101 N. Elm, Fargo, ND 58102

- VA Health Care Upstate New York (James Norton, Ph.D.)

113 Holland Avenue, Albany, NY 12208

- New Mexico VA Health Care System (Gerardo Villareal, M.D.)

1501 San Pedro Drive, S.E. Albuquerque, NM 87108

- VA Boston Healthcare System (Scott Kinlay, M.B.B.S., Ph.D.)

150 S. Huntington Avenue, Boston, MA 02130

- VA Western New York Healthcare System (Junzhe Xu, M.D.)

3495 Bailey Avenue, Buffalo, NY 14215-1199

- Ralph H. Johnson VA Medical Center (Mark Hamner, M.D.)

109 Bee Street, Mental Health Research, Charleston, SC 29401

- Columbia VA Health Care System (Roy Mathew, M.D.)

6439 Garners Ferry Road, Columbia, SC 29209

- VA North Texas Health Care System (Sujata Bhushan, M.D.)

4500 S. Lancaster Road, Dallas, TX 75216

- Hampton VA Medical Center (Pran Iruvanti, D.O., Ph.D.)

100 Emancipation Drive, Hampton, VA 23667

- Richmond VA Medical Center (Michael Godschalk, M.D.)

1201 Broad Rock Blvd., Richmond, VA 23249

- Iowa City VA Health Care System (Zuhair Ballas, M.D.)

601 Highway 6 West, Iowa City, IA 52246-2208

- Eastern Oklahoma VA Health Care System (River Smith, Ph.D.)

1011 Honor Heights Drive, Muskogee, OK 74401

- James A. Haley Veterans’ Hospital (Stephen Mastorides, M.D.)

13000 Bruce B. Downs Blvd, Tampa, FL 33612

- James H. Quillen VA Medical Center (Jonathan Moorman, M.D., Ph.D.)

Corner of Lamont & Veterans Way, Mountain Home, TN 37684

- John D. Dingell VA Medical Center (Saib Gappy, M.D.)

4646 John R Street, Detroit, MI 48201

- Louisville VA Medical Center (Jon Klein, M.D., Ph.D.)

800 Zorn Avenue, Louisville, KY 40206

- Manchester VA Medical Center (Nora Ratcliffe, M.D.)

718 Smyth Road, Manchester, NH 03104

- Miami VA Health Care System (Ana Palacio, M.D., M.P.H.)

1201 NW 16th Street, 11 GRC, Miami FL 33125

- Michael E. DeBakey VA Medical Center (Olaoluwa Okusaga, M.D.)

2002 Holcombe Blvd, Houston, TX 77030

- Minneapolis VA Health Care System (Maureen Murdoch, M.D., M.P.H.)

One Veterans Drive, Minneapolis, MN 55417

- N. FL/S. GA Veterans Health System (Peruvemba Sriram, M.D.)

1601 SW Archer Road, Gainesville, FL 32608

- Northport VA Medical Center (Shing Shing Yeh, Ph.D., M.D.)

79 Middleville Road, Northport, NY 11768

- Overton Brooks VA Medical Center (Neeraj Tandon, M.D.)

510 East Stoner Ave, Shreveport, LA 71101

- Philadelphia VA Medical Center (Darshana Jhala, M.D.)

3900 Woodland Avenue, Philadelphia, PA 19104

- Phoenix VA Health Care System (Samuel Aguayo, M.D.)

650 E. Indian School Road, Phoenix, AZ 85012

- Portland VA Medical Center (David Cohen, M.D.)

3710 SW U.S. Veterans Hospital Road, Portland, OR 97239

- Providence VA Medical Center (Satish Sharma, M.D.)

830 Chalkstone Avenue, Providence, RI 02908

- Richard Roudebush VA Medical Center (Suthat Liangpunsakul, M.D., M.P.H.)

1481 West 10th Street, Indianapolis, IN 46202

- Salem VA Medical Center (Kris Ann Oursler, M.D.)

1970 Roanoke Blvd, Salem, VA 24153

- San Francisco VA Health Care System (Mary Whooley, M.D.)

4150 Clement Street, San Francisco, CA 94121

- South Texas Veterans Health Care System (Sunil Ahuja, M.D.)

7400 Merton Minter Boulevard, San Antonio, TX 78229

- Southeast Louisiana Veterans Health Care System (Joseph Constans, Ph.D.)

2400 Canal Street, New Orleans, LA 70119

- Southern Arizona VA Health Care System (Paul Meyer, M.D., Ph.D.)

3601 S 6th Avenue, Tucson, AZ 85723

- Sioux Falls VA Health Care System (Jennifer Greco, M.D.)

2501 W 22nd Street, Sioux Falls, SD 57105

- St. Louis VA Health Care System (Michael Rauchman, M.D.)

915 North Grand Blvd, St. Louis, MO 63106

- Syracuse VA Medical Center (Richard Servatius, Ph.D.)

800 Irving Avenue, Syracuse, NY 13210

- VA Eastern Kansas Health Care System (Melinda Gaddy, Ph.D.)

4101 S 4th Street Trafficway, Leavenworth, KS 66048

- VA Greater Los Angeles Health Care System (Agnes Wallbom, M.D., M.S.)

11301 Wilshire Blvd, Los Angeles, CA 90073

- VA Long Beach Healthcare System (Timothy Morgan, M.D.)

5901 East 7th Street Long Beach, CA 90822

- VA Maine Healthcare System (Todd Stapley, D.O.)

1 VA Center, Augusta, ME 04330

- VA New York Harbor Healthcare System (Peter Liang, M.D., M.P.H.)

423 East 23rd Street, New York, NY 10010

- VA Pacific Islands Health Care System (Daryl Fujii, Ph.D.)

459 Patterson Rd, Honolulu, HI 96819

- VA Palo Alto Health Care System (Philip Tsao, Ph.D.)

3801 Miranda Avenue, Palo Alto, CA 94304-1290

- VA Pittsburgh Health Care System (Patrick Strollo, Jr., M.D.)

University Drive, Pittsburgh, PA 15240

- VA Puget Sound Health Care System (Edward Boyko, M.D.)

1660 S. Columbian Way, Seattle, WA 98108-1597

- VA Salt Lake City Health Care System (Jessica Walsh, M.D.)

500 Foothill Drive, Salt Lake City, UT 84148

- VA San Diego Healthcare System (Samir Gupta, M.D., M.S.C.S.)

3350 La Jolla Village Drive, San Diego, CA 92161

- VA Sierra Nevada Health Care System (Mostaqul Huq, Pharm.D., Ph.D.)

975 Kirman Avenue, Reno, NV 89502

- VA Southern Nevada Healthcare System (Joseph Fayad, M.D.)

6900 North Pecos Road, North Las Vegas, NV 89086

- VA Tennessee Valley Healthcare System (Adriana Hung, M.D., M.P.H.)

1310 24th Avenue, South Nashville, TN 37212

- Washington DC VA Medical Center (Jack Lichy, M.D., Ph.D.)

50 Irving St, Washington, D. C. 20422

- W.G. (Bill) Hefner VA Medical Center (Robin Hurley, M.D.)

1601 Brenner Ave, Salisbury, NC 28144

- White River Junction VA Medical Center (Brooks Robey, M.D.)

163 Veterans Drive, White River Junction, VT 05009

- William S. Middleton Memorial Veterans Hospital (Prakash Balasubramanian, M.D.)

2500 Overlook Terrace, Madison, WI 53705

**Supplementary Tables**

**Supplementary Table 1. Genome-wide significant variants from the GBMI stroke meta-analysis.** For all variants the effect sizes are reported for the minor allele.

| rsID | Chr | POS (hg38) | EA/  NEA | EAF | N samples | Beta  (SE) | P-value | Sex-  heterogeneity  Q stat | Sex-  heterogeneity P-value | Annotation  Closest gene(s) |
| --- | --- | --- | --- | --- | --- | --- | --- | --- | --- | --- |
| Putative novel loci | | | | | | | | | | |
| *rs201194999 | 4 | 65801177 | T/C | 0.0475 | 443,083 | 0.183  (0.029) | 3.48E-10 | 5.96 | 0.015 | Intergenic  (EPHA5-AS1/MIR1269A) |
| rs12509595 | 4 | 80261400 | C/T | 0.308 | 1,319,212 | 0.0474  (0.0073) | 6.66E-11 | 1.55 | 0.213 | Intergenic  (PRDM8/FGF5) |
| rs3131618 | 6 | 31466844 | G/A | 0.120 | 995,658 | 0.0965  (0.016) | 5.93E-10 | 0.636 | 0.425 | Intergenic  (HLA-region, HLA-C) |
| rs2501968 | 6 | 49492265 | G/A | 0.446 | 1,320,046 | -0.0457  (0.0068) | 1.80E-11 | 1.96 | 0.162 | Exonic  CENPQ |
| *rs111910126 | 7 | 41566524 | T/C | 0.0671 | 540,497 | -0.277  (0.043) | 7.78E-11 | 5.48 | 0.019 | Intergenic  (LINC01449/INHBA) |
| NA | 12 | 89687411 | C/T | 0.618 | 1,297,864 | 0.0383  (0.0070) | 3.71E-08 | 2.36 | 0.124 | Intronic  (ATP2B1) |
| rs146765815 | 15 | 37490789 | T/G | 4.88E-3 | 5,156 | 3.34  (0.61) | 4.01E-08 | NA** | NA** | Intergenic  (MEIS2/TMCO5A) |
| Previously published loci | | | | | | | | | | |
| rs1275985 | 2 | 26688877 | T/C | 0.515 | 1,319,173 | -0.0439  (0.0072) | 1.20E-09 | 0.292 | 0.589 | Intergenic  (CIB4/KCNK3) |
| rs1333047 | 9 | 22124505 | T/A | 0.519 | 1,318,621 | 0.0613  (0.0069) | 7.26E-19 | 0.761 | 0.383 | Intergenic  (CDKN2B-AS1/DMRTA1) |
| rs7091346 | 10 | 103849066 | T/C | 0.429 | 1,138,816 | -0.0467  (0.0082) | 1.33E-08 | 92.6 | 6.27E-22 | Intronic  (SH3PXD2A) |
| rs12811752 | 12 | 20424871 | C/T | 0.455 | 1,320,000 | 0.0496  (0.0068) | 4.25E-13 | 0.398 | 0.528 | Intronic  (PDE3A) |
| rs671 | 12 | 111803962 | A/G | 0.238 | 293,460 | -0.107  (0.012) | 5.16E-18 | 25.6 | 4.15E-07 | Exonic  (ALDH2) |
| rs4773140 | 13 | 110301890 | G/A | 0.740 | 1,155,606 | 0.0489  (0.0081) | 1.28E-09 | 3.92 | 0.048 | Intronic  (COL4A1) |
| rs11880613 | 19 | 10793296 | A/G | 0.173 | 1,318,912 | -0.0508  (0.0089) | 1.00E-08 | 0.734 | 0.392 | Intronic  (DNM2) |

* Driven by a single cohort in the GBMI discovery

** Sex specific summary statistics not available due to low number of variant carriers in the contributing cohorts

Chr = Chromosome; POS = Position; EA = Effect allele; NEA = Non-effect allele; N = Number of samples; SE = Standard error of the effect size; Q-stat = Cochran’s Q-statistic

**Supplementary Table 2. Replication results for all 7 putative novel loci.** The GBMI discovery is based on the leave BioMe out to correct for the fact that BioMe is included in the TOPMed replication results. (Accompanying excel)

**Supplementary Table 3A. Significant expression quantitative trait loci (eQTL) association results for the two novel lead SNPs** (Accompanying excel)

**Supplementary Table 3B. Significant splicing quantitative trait loci (sQTL) association results for the two novel lead SNPs** (Accompanying excel)

**Supplementary Table 4. Sex-specific association results for loci showing significant sex-heterogeneity.** For all variants the effect sizes are reported for the minor allele.

| **Variant information** | | | | **Males** | | | **Females** | | |
| --- | --- | --- | --- | --- | --- | --- | --- | --- | --- |
| **rsID** | **Chromosome** | **Position (hg38)** | **EA/ NEA** | **Effect size (SE)** | **N cases /**  **N controls** | **P-value** | **Effect size (SE)** | **N cases /**  **N controls** | **P-value** |
| rs7091346 | 10 | 103849066 | T/C | -0.062 (0.011) | 20673 / 404273 | 4.84e-8 | -0.031 (0.014) | 14426 / 479632 | 0.022 |
| rs671 | 12 | 111803962 | A/G | -0.160 90.016) | 14186 / 80046 | 1.67e-24 | -0.031 (0.020) | 8702 /  87238 | 0.126 |

EA = Effect allele; NEA = Non-effect allele, SE = Standard error of the effect size; N = number of samples

**Supplementary Figures**

**Supplementary Figures 1A-B. Locus zoom plots for the newly identified loci from the GBMI multi-ancestry analysis.** Y-axis shows the -log10(P-value) and the X-axis shows the genome position in hg19. The linkage disequilibrium is shown in reference to the locus lead variant.

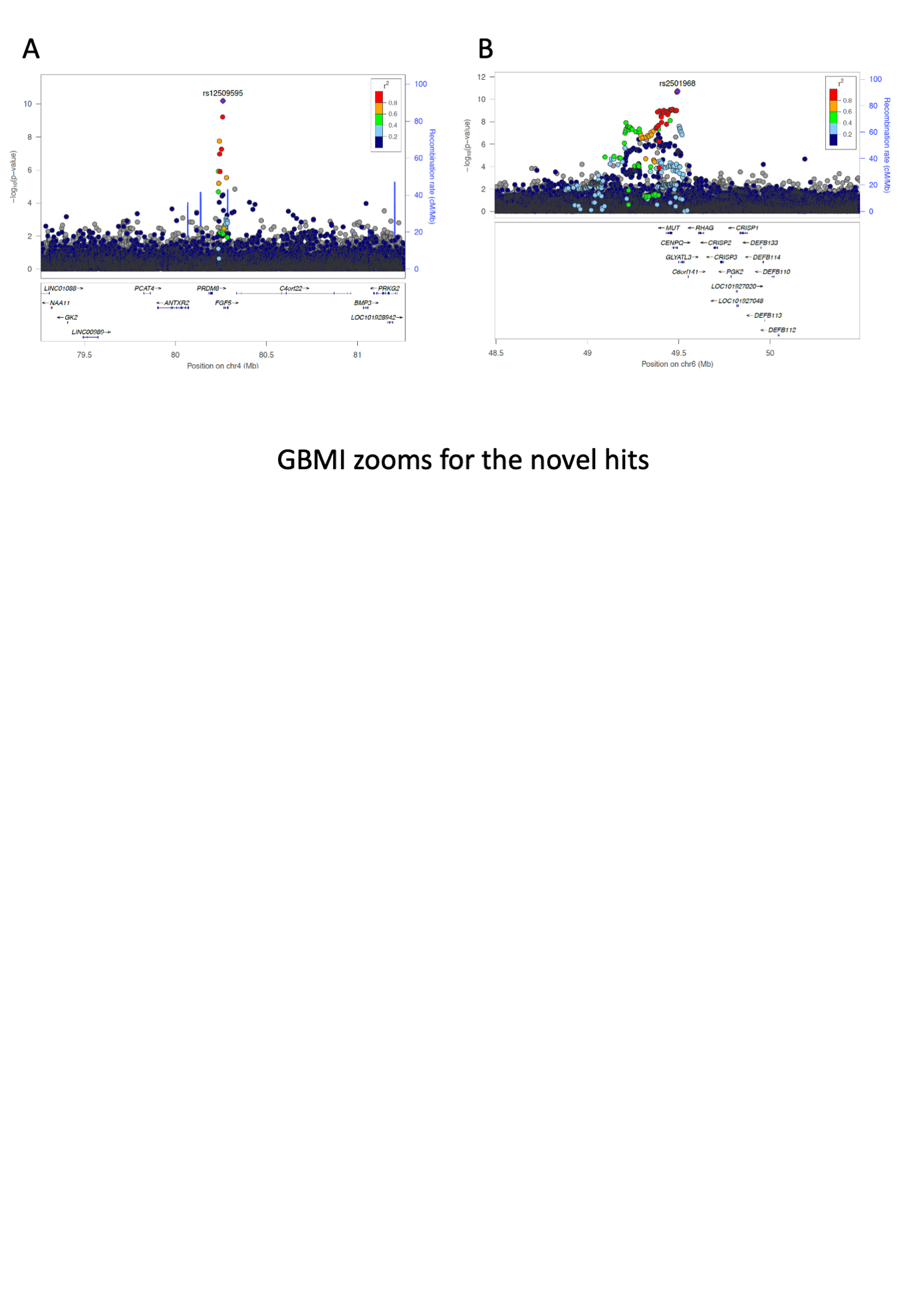

**Supplementary Figures 2A-B. Locus zoom plots for the newly identified loci from the MegaStroke Consortium.** Y-axis shows the -log10(P-value) and the X-axis shows the genome position in hg19. The linkage disequilibrium is shown in reference to the most significant variant in the locus in MegaStroke summary statistics.

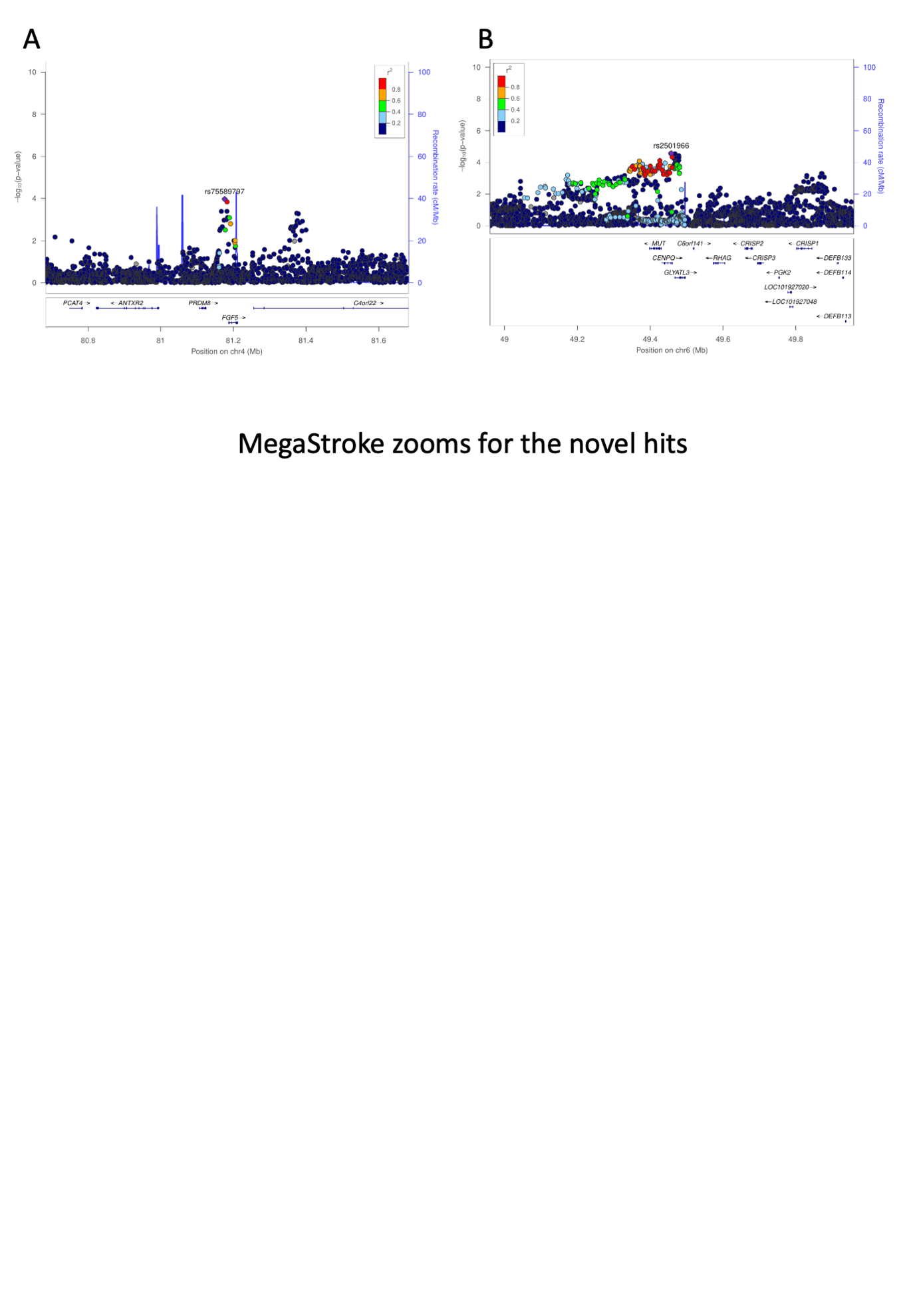

**Supplementary Figure 3A-B. Forest plots for the lead variants showing significant sex-heterogeneity.** Panel A shows the results for the *SH3PXD2A* locus in chromosome 10 and panel B for *ALDH2* locus in chromosome 12. The x-axis represents the effect size and 95% confidence intervals are shown with the whiskers.

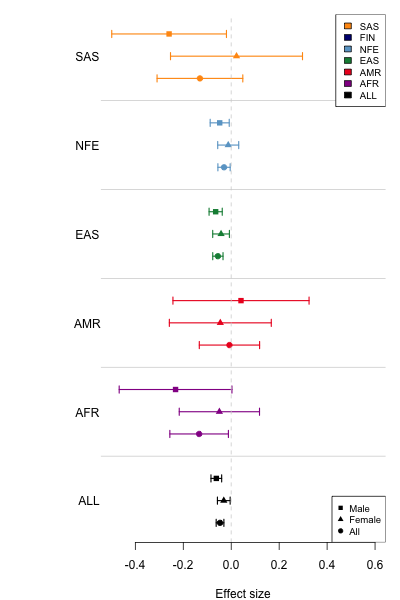

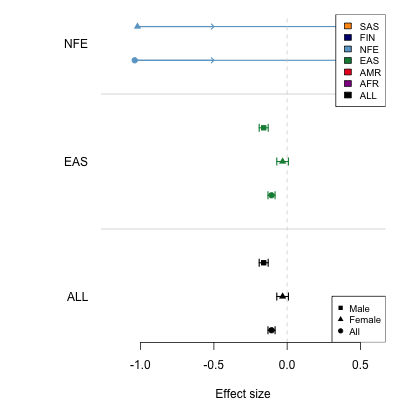

**Supplementary Figure 4. C-index for different stroke Cox models in HUNT.** Presented are the C-indexes for the reference model with age and sex only for the sex-combined (all), males and females and for the models where either PRS based on the sex-combined summary statistics (joint score) or based on the sex-specific summary statistics (male/female score) was added on top of the age and sex.

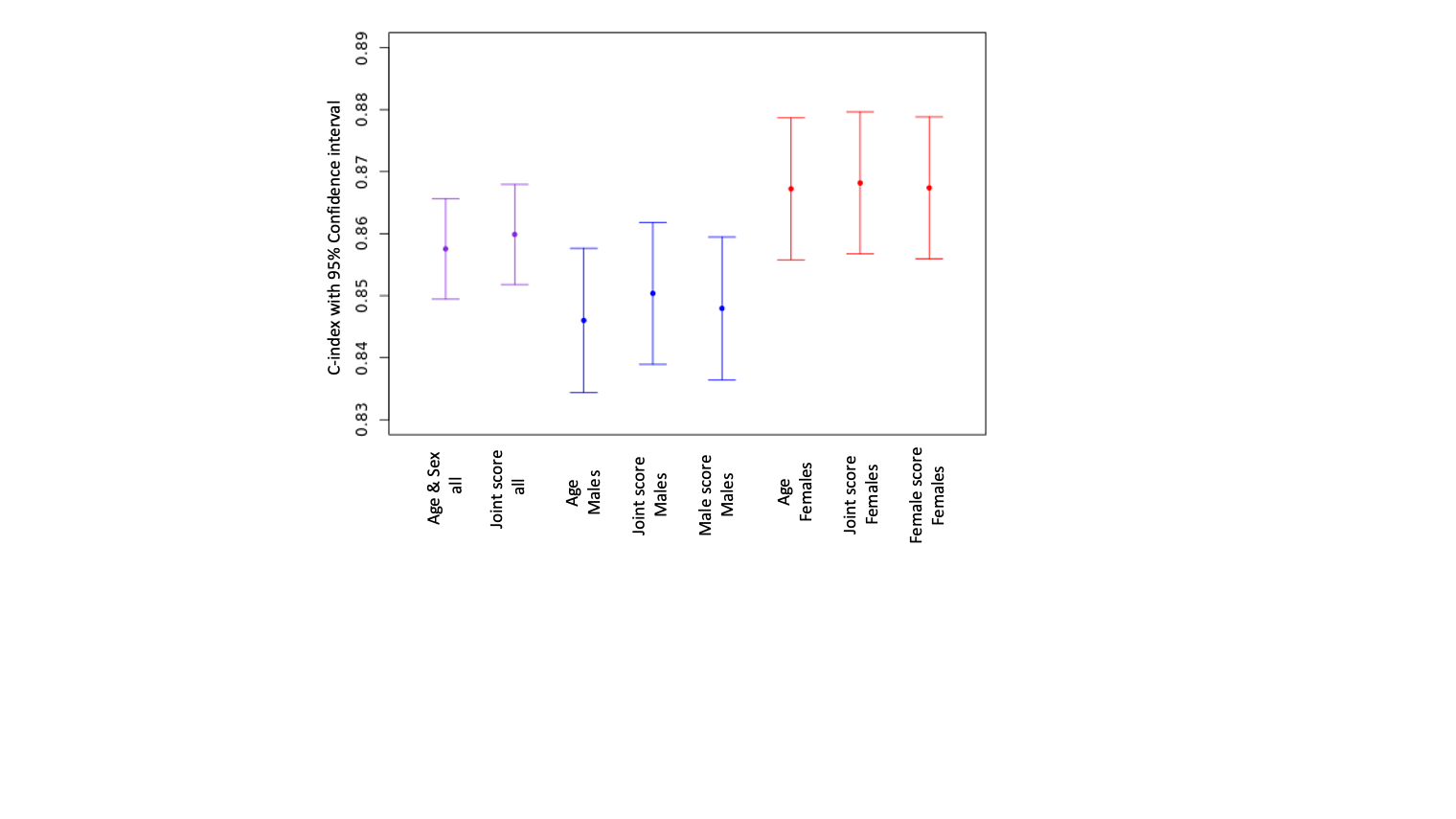
